# Real-time situational assessment of respiratory virus epidemics in Australia over winter 2025 and an out-of-season influenza A H3N2 epidemic

**DOI:** 10.64898/2026.09.30.26364449

**Authors:** Oliver Eales, Alys R. Young, Katharine L. Senior, Ruarai Tobin, Janaki Amin, Brett Nicholas Archer, Paul Armstrong, Sally Ellis, Elizabeth J. Robinson, Janet Strachan, Ashish Chandra Shrestha, Mark Veitch, James Wood, James M McCaw, Freya M. Shearer

## Abstract

**Introduction:** In temperate regions, influenza and respiratory syncytial virus (RSV) typically exhibit seasonal epidemics during the winter months, often concurrently with SARS-CoV-2 epidemics. Monitoring past and real-time trends in infection dynamics is important for informing public health situational awareness, including identifying and characterising unusual epidemic activity. However, data used to monitor these trends can exhibit substantial noise including strong day-of-the-week variation in reporting. In addition, case data for influenza can obscure distinct trends in influenza subtypes if not combined with virological data.

**Aim:** By quantifying trends in respiratory virus activity using statistical methods, the Australia-Aotearoa Consortium for Epidemic Forecasting and Analytics (ACEFA) aims to provide government partners with comprehensive epidemic intelligence.

**Methods:** Analyses were performed on daily case time series data for SARS-CoV-2, RSV and influenza across Australia’s eight jurisdictions. Past and real-time trends were inferred for each pathogen and each influenza subtype using Bayesian p-spline models.

**Results:** The 2025 Winter Situational Assessment Program provided weekly reports throughout the winter season and during the out-of-season influenza A H3N2 epidemic, driven by the emergence of the novel influenza subclade (‘subclade K’). Despite limited subtyping data, our analysis provided an early signal of increasing influenza A H3N2 during a period in which overall influenza cases were decreasing; increases in influenza A H3N2 activity were first detected in July, and by mid-August, increasing H3N2 activity was estimated across all jurisdictions.

**Discussion:** Our analysis highlights the benefits of incorporating statistical methods into real-time epidemic situational assessment and in consolidating reporting across jurisdictions.

## Introduction

Many respiratory viruses, including influenza and respiratory syncytial virus (RSV), exhibit seasonal transmission dynamics, with clinical burden typically peaking over winter months in temperate regions of the southern and northern hemispheres [1,2]. In tropical and subtropical regions, seasonality is less distinct and multiple peaks may be observed or periods of sustained (often year-round) activity. Meanwhile, it is unclear if SARS-CoV-2 will settle into a regular epidemic cycle, but increased activity was observed during winter 2025.

Monitoring past and real-time trends in infection levels is important for informing public health situational awareness and potential response measures. By analysing the past seasonal dynamics of respiratory pathogens, we can improve predictions of season onset, peak timing and peak burden of future seasons [3]. Furthermore, knowledge of typical seasonal dynamics, when combined with real-time analysis, can help to rapidly identify and characterise unusual epidemic activity. For example, at the end of 2025 (and early-2026), influenza activity continued unusually late in the southern hemisphere due to emergence of a novel subclade of influenza A H3N2 (‘subclade K’) [4]. This was paralleled by an early and rapid start to the 2025/2026 season in the northern hemisphere [5,6].

Case-based surveillance systems are an important data source for monitoring trends in respiratory pathogen infections [7]. However, in addition to biases caused by time-varying reporting rates [7], case time series can exhibit substantial noise and day-of-the-week reporting variations, which can make it difficult to interpret trends in the data. Statistical models can infer smooth trends (including uncertainty) in time series case data, by accounting for noise and day-of-the-week effects [8,9]. From smooth trends, quantities such as growth rates, doubling/halving times, and the probability that cases are increasing, stable, or decreasing can be estimated and reported to support situational awareness.

Many influenza surveillance systems also record influenza type/subtype (e.g. influenza A H1N1, influenza AH3N2, and influenza B) for a subset of cases that have undergone additional virological testing. While these data are often sparse and noisy (e.g. 4% of influenza A cases were subtyped in Australia in 2025), they are critical for understanding seasonal influenza dynamics which are frequently caused by concurrent circulation of multiple subtypes, each with distinct temporal dynamics. Inferring trends in influenza cases, under the assumption that these trends correlate with the infection levels of only a single subtype, can lead to misleading conclusions [10]. For example, if two influenza subtypes are circulating and the incidence of one subtype is decreasing from high prevalence while that of another is increasing from low prevalence, analysing the combined case dynamics would obscure the emergence of the latter subtype. This was the situation when influenza A H3N2 subclade K emerged in Australia in mid-2025. However, statistical methods can be used to jointly analyse influenza case time series data and subtyping data, inferring the individual dynamics of each subtype and providing more comprehensive epidemic intelligence [10].

Here we present end of season and real-time estimates of trends in case dynamics of SARS-CoV-2, RSV, influenza, and influenza types/subtypes across all eight Australian jurisdictions for 2025, generated as part of the Australia-Aotearoa Consortium for Epidemic Forecasting and Analytics (ACEFA) Winter Situational Assessment Program. Real-time estimates were produced over the regular respiratory pathogen season (22 weeks of reporting, 16 May-3 October 2025) and for influenza only over an inter-seasonal period due to the late influenza A H3N2 epidemic (nine weeks of reporting, 6 November 2025-4 January 2026).

## Methods

### Data

This study received research ethics approval from the University of Melbourne Human Research Ethics Committee (application identifier 2024-26949-50575-3).

Data was provided weekly by the National Notifiable Diseases Surveillance System (NNDSS) and the Victorian Government Department of Health. The latter source was used for all Victorian (VIC) data during the study period, while NNDSS data was used for the remaining seven states and/or territories: the Australian Capital Territory (ACT), New South Wales (NSW), the Northern Territory (NT), Queensland (QLD), South Australia (SA), Tasmania (TAS) and Western Australia (WA). We received de-identified case data in linelist format and aggregated this to daily counts of laboratory-confirmed cases by notification date for each jurisdiction and pathogen (SARS-CoV-2 [11], RSV [12] and influenza [13]). For influenza, daily case counts by notification date (for each jurisdiction) were also aggregated by type and subtype. Influenza type information was available for approximately 99% of influenza cases, while subtyping information was unavailable (ACT, SA) or only available for between 0.6% (VIC) and 18.4% (TAS) of cases over 2025 (Table S1). For Victorian data only, we filtered data for all pathogens to include only cases reported by electronic laboratory reporting (ELR) to exclude known delays in other reporting formats [18]. Across 2025, ELR cases represented 100%, 89.6% and 87.7% of total cases of SARS-CoV-2, RSV, and influenza, respectively. Information on the reporting method was not available for data supplied by NNDSS, so this exclusion was not applied to other jurisdictions.

Although data was provided on a consistent day each week, the final day of data was not consistent between jurisdictions, pathogens or reporting week. We removed any cases dated the same day as data was received, as data was received in the middle of the day and would miss any cases reported after that point.

### Statistical models

We used a single-pathogen Bayesian P-spline model [8] to infer temporal trends of RSV and SARS-CoV-2 in each jurisdiction. The model takes as input a time-series of daily cases. The statistical model infers smooth trends in cases over time accounting for noise and day-of-the-week effects in case reporting.

For influenza, type/subtype data were incorporated where available, using a multi-pathogen Bayesian P-spline model [8] to infer distinct trends of: influenza types (influenza A and influenza B) in the ACT and SA where subtype data were not available; and influenza types and subtypes (influenza A H1N1, influenza A H3N2, and influenza B) in all other jurisdictions. The trends in overall influenza cases were presented by summing the inferred trends in all types/subtypes. The model takes as input: (1) a time-series of the daily number of overall influenza cases; and (2) a time-series of type/subtype data describing the number of influenza cases that were determined to be influenza A (subtype not determined), influenza A H3N2, influenza A H1N1, and influenza B. The statistical model infers smooth trends in cases attributable to each influenza type/subtype and overall (the sum of individual subtype contributions) accounting for noise, day-of-the-week effects, typing/subtyping rates that vary over time, and influenza A samples that are not subtyped (i.e. influenza A with subtype not determined). For the single-pathogen and multi-pathogen Bayesian P-spline models we used the default settings described in [8] (i.e. third order basis splines, five days between adjacent knots, independent smoothness parameter for each influenza subtype)

We inferred and reported the smoothed trends in reported cases (as opposed to infections). From the smoothed trends we also calculated the epidemic growth rate (*r*), the probability the growth rate is greater than 0 (i.e. the probability that cases are increasing), and the doubling/halving time. The doubling/halving time is the time taken for cases to reach double/half of their current value if the growth rate remains constant at its current value (doubling/halving time = *ln*(2)/*r*).

### Retrospective analysis

We fitted statistical models to data received from NNDSS and the Victorian Government Department of Health on 14 January 2026. For each pathogen and jurisdiction combination, we fit the model to all data available from 1 January 2023 (SFig.1-SFig.10) up to the penultimate day of full data included in the final dataset (due to partial reporting).

### Real-time analysis

We fitted statistical models in real-time to data received weekly from 16 May-3 October 2025 (22 rounds, regular season reports) and additionally for influenza from 6 November-25 December 2025, and on 14 January 2026 following receipt of the final data set (9 rounds, inter-season reports), due to the late influenza A H3N2 epidemic. For each pathogen and jurisdiction combination, we fit the real-time model to the previous three years of data up to the last day of data. The last day of data included in the model was usually the last full day of data. However, data was cut off earlier for some jurisdictions/rounds as a result of known/pre-identified delays in reporting and hence consistent revisions upwards of the last day(s) in the following week’s data. This was done for SARS-CoV-2 in WA from 18 June onward (last two full days of data removed) and for RSV and influenza in WA from 24 September (last full day of data removed) onward. Additionally, for inter-season reports of influenza activity (nine rounds) an additional day of data was removed for all jurisdictions, as the last day of data was retrospectively found to have been frequently revised during the regular winter reporting (not only in WA). Real-time estimates were only made for SA from 10 July 2025 (report 9) onwards when SA data became available for analysis. Influenza type specific models (influenza A and influenza B) for the ACT and SA were only performed from 10 July 2025 (report 9) onwards. All real-time estimates were those presented in ACEFA’s weekly situational assessments provided to government partners. We compared real-time estimates to end of season estimates to assess how models performed over the season, particularly when there were significant data revisions.

## Results

### Retrospective analyses

#### SARS-CoV-2

COVID-19 cases began increasing (central estimate of the growth rate greater than zero) across all eight jurisdictions from approximately April 2025 (Fig.1). Cases increased until approximately June 2025, when epidemic activity peaked and began to decrease. In SA, QLD, and VIC, transient increases were observed in the growth rate of SARS-CoV-2 as early as March 2025 (SFig.11); this resulted in a small increase (SA, QLD) or stabilisation (VIC) of cases prior to the primary epidemic. In the NT, VIC, and WA, transient increases in the growth rate occurred later in the season from July to early-August, after the epidemics had peaked. This resulted in a second epidemic peak in the NT, and a period of stable epidemic activity in VIC and WA.

**Figure 1:**
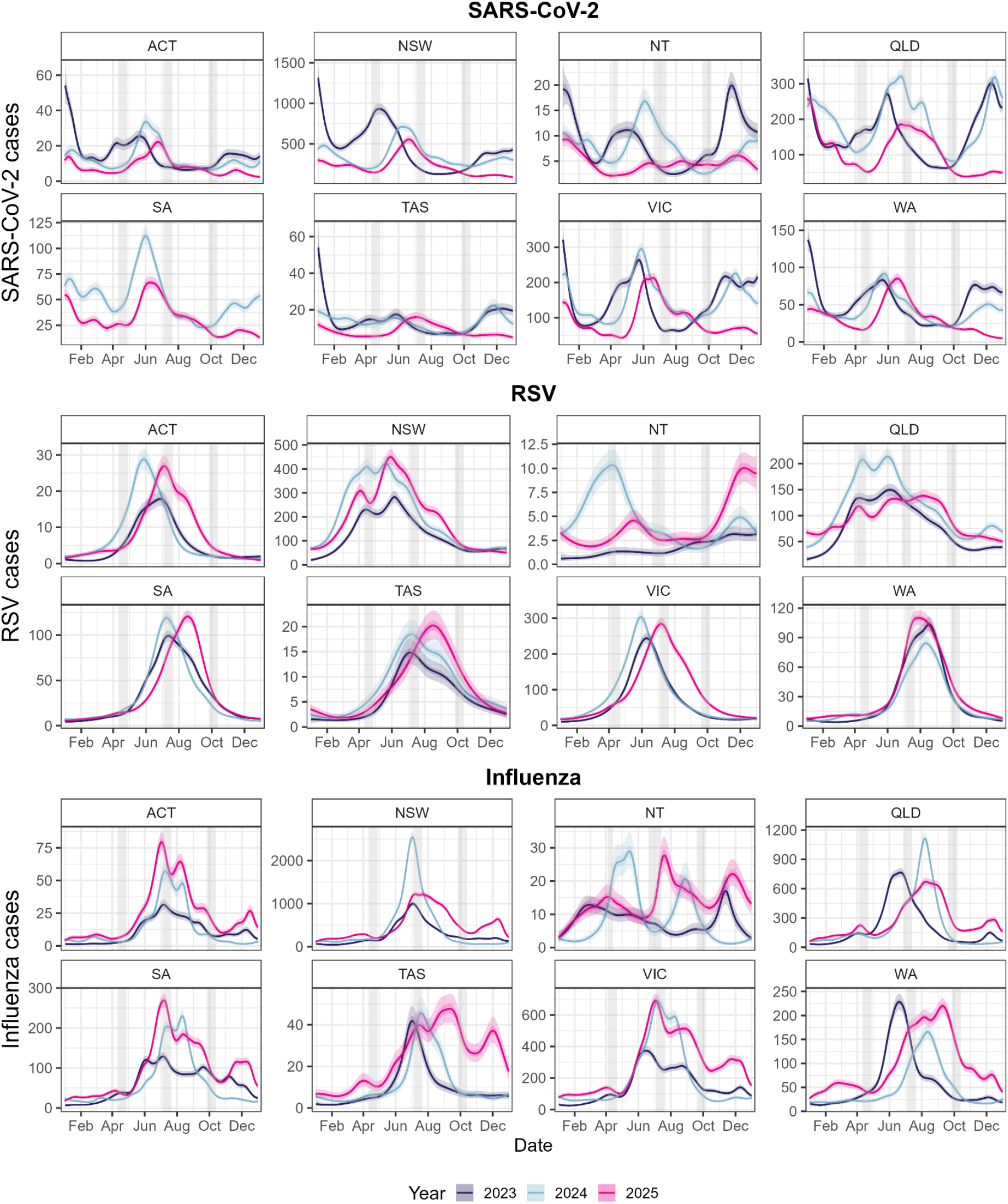
Comparison of case dynamics of SARS-CoV-2, RSV, and influenza in 2023, 2024 and 2025. Modelled estimates of the expected number of SARS-CoV-2, RSV, and influenza cases over time for 2025 (pink), 2024 (light blue), and 2023 (dark blue). All estimates are shown over time with median (line), 50% credible intervals (dark shaded region) and 95% credible intervals (light shaded region). Comparison of modelled estimates to data are provided in Supplementary Figures 1-8. The grey shaded regions highlight the Autumn, Winter and Spring school holidays for each jurisdiction in 2025. Note that SARS-CoV-2 data were not available for SA for 2023.

In all jurisdictions, except TAS and the NT, SARS-CoV-2 case dynamics from January to October were broadly similar to the 2024 season, but with lower peak magnitude and slightly later peak timing (Fig.1, SFig.11). The difference in peak timing between the corresponding seasons in each year ranged from (approximately) no difference (QLD) to five weeks (WA). The timing of epidemics was also broadly similar to the 2023 season but occurring even later (e.g. up to nine weeks later in NSW). In all jurisdictions SARS-CoV-2 cases were low from October onwards, relative to the same period in 2023 and 2024 in which there were large epidemics — peaking in approximately November/December (i.e. Australian summer). Despite the low level of epidemic activity, in all jurisdictions there were transient increases in the growth rate in late-2025 with similar timing to the 2024 season (SFig.11); this led to small resurgences in SARS-CoV-2 cases.

#### RSV

We estimated that RSV cases began increasing (central estimate of the growth rate greater than zero) from as early as December 2024 (ACT, SA, WA), and from as late as early-March 2025 (NT, TAS) (SFig.12). RSV epidemics were longer in duration relative to SARS-CoV-2, with earlier onset in all jurisdictions and later peak timing in all jurisdictions except NSW and the NT (Fig.1). In NSW and the NT, RSV peaked in May (before SARS-CoV-2), whereas in all other jurisdictions peaked in July or August (after SARS-CoV-2).

While the magnitude of the epidemics varied, RSV case dynamics were similar to both the 2023 and 2024 seasons in all jurisdictions except the NT (Fig.1, SFig.12). In the ACT, SA, TAS, VIC and WA, single epidemic peaks were observed for all three seasons, with peaks slightly later in the 2025 season for the ACT, SA, TAS, and VIC (WA had comparable timing between seasons). In NSW and QLD, two RSV peaks were observed during the 2023 and 2024 seasons. In NSW a double peak pattern was also observed during the 2025 season with similar timing to previous seasons (peaks in approximately early-April and late-May). In QLD the double peak pattern was less apparent in 2025, with the second peak prolonged over several months (and potentially including a third peak). However, trends in the growth rate for QLD were similar between seasons, decreasing after the second peak (but remaining approximately zero) in line with patterns in 2023 and 2024.

#### Influenza/influenza subtypes

Influenza cases began increasing (central estimate of the growth rate greater than zero) across all eight jurisdictions in late-2024, leading to early-season epidemics that peaked between early-March and early-April 2025 (Fig.1, SFig.13). Following these early season epidemics, influenza cases began increasing approximately one month later (between early-April and early-May) in most jurisdictions — except in the NT, where there was significant uncertainty as to when cases began increasing (SFig.13). Sharp decreases in the epidemic growth rate were estimated in late-June/early-July across all jurisdictions (except the NT), coinciding (approximately) with the beginning of school holidays (timing differed between jurisdictions); this resulted in an epidemic peak in the ACT, NSW, SA,TAS and VIC. In some jurisdictions (ACT, SA, VIC) a rebound in the growth rate occurred from late-July, coinciding (approximately) with the end of school holidays.

A high degree of heterogeneity in influenza cases trends was observed between seasons for some jurisdictions (Fig.1). For example, in WA influenza cases peaked approximately 4 and 10 weeks later in 2025 compared to 2024 and 2023 respectively. In the NT, epidemics peaked at different times in 2023 (late-Feb and mid-November), 2024 (late-May and late-August), and 2025 (early-April and mid-July). In other jurisdictions, trends in influenza cases were similar across seasons. In VIC, similar trends in the epidemic growth rate were observed in the 2023, 2024, and 2025 season, and similar patterns in cases; an epidemic peak around late-June/early-July followed by a temporary decrease and secondary (lower) peak in August.

Our subtype specific model showed that influenza A H1N1 was the predominant influenza subtype for most of the period from January-September 2025, followed by influenza B, across all jurisdictions for which subtype data were available to the consortium (Fig.2, Fig.3, SFig.14). While there was limited influenza A H3N2 activity early in the season, our multi-pathogen model estimated that cases began increasing from approximately May to June, resulting in unusually late and large epidemics that peaked from late-November to mid-December (summer). Transient decreases in the growth rate of influenza A H3N2 were observed in late-September/early-October, aligning with spring school holidays (Fig.3). In the ACT and SA, where subtype data were not available, influenza A was the predominant type for most of the season. In both jurisdictions, increases in the epidemic growth rate of influenza A were estimated in mid-August, which likely reflected increasing H3N2 activity.

**Figure 2:**
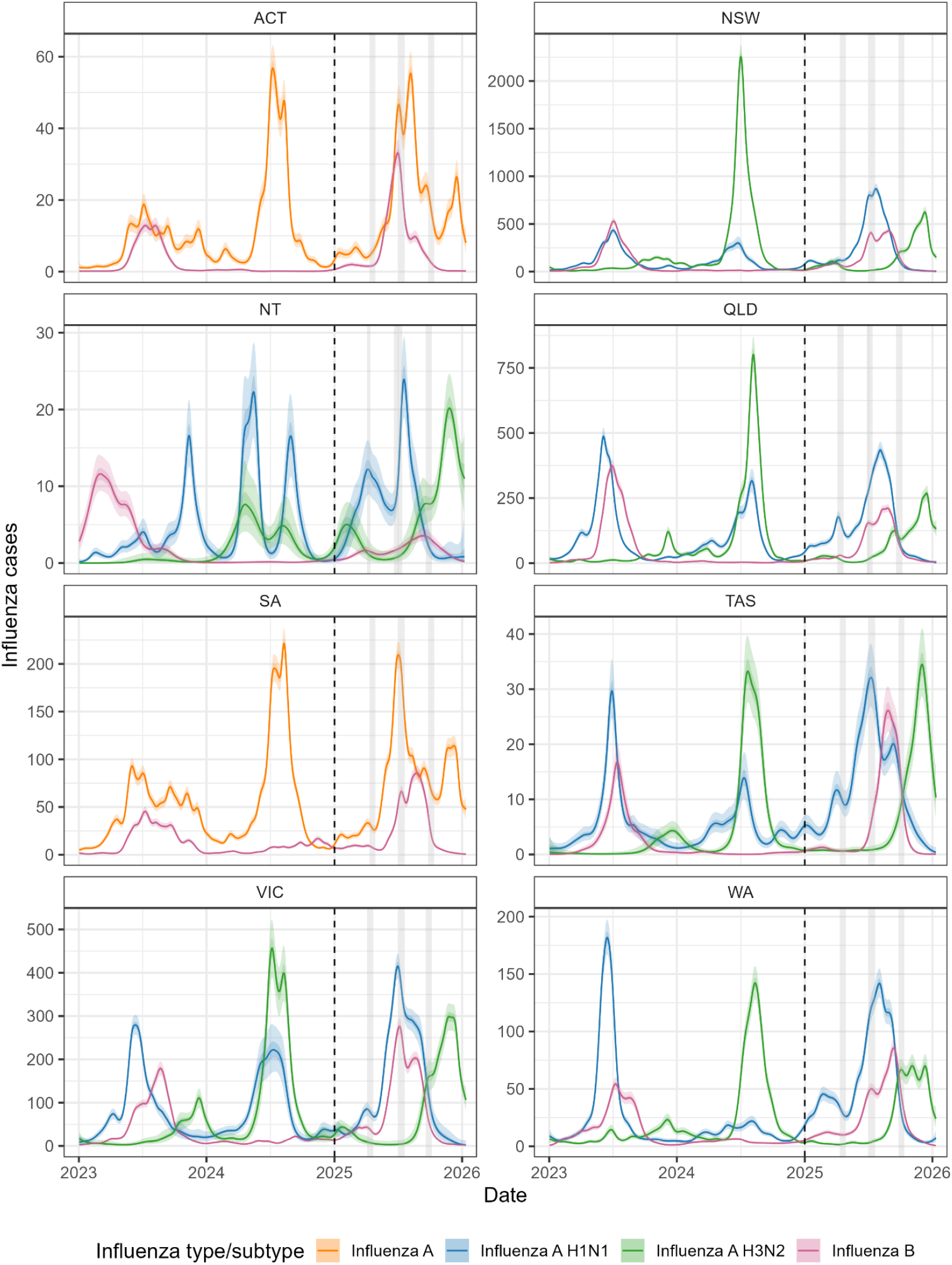
Modelled case dynamics of influenza subtypes over the previous three seasons. Modelled estimates of the expected number of cases over time for each influenza subtype. All estimates are shown over time with median (line), 50% credible intervals (dark shaded region) and 95% credible intervals (light shaded region). Comparison of the modelled relative proportions of each subtype compared to data are provided in Supplementary Figures 9-10. The vertical dashed line highlights the start of the 2025 season which is the focus of this paper. The grey shaded regions highlight the Autumn, Winter and Spring school holidays for each jurisdiction in 2025. Overall trends in influenza A for all jurisdictions are provided in Supplementary Figure 26 for visual comparison to trends in influenza A in the ACT and SA.

**Figure 3:**
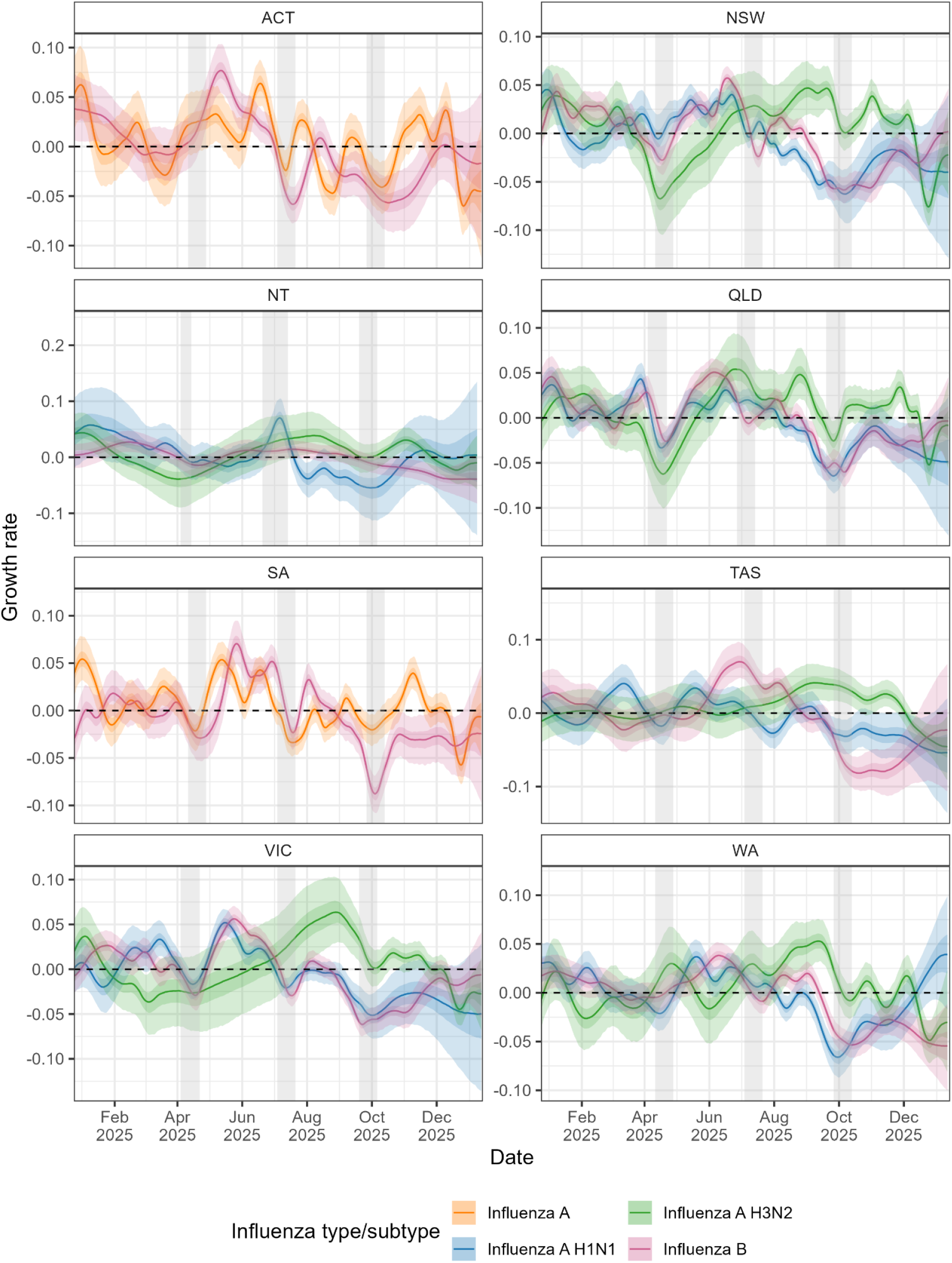
Modelled growth rate of influenza subtypes over the 2025 season. Modelled estimates of the daily growth rate for each influenza subtype. The dashed line shows where growth rate equals 0, the transition point between epidemic growth and decline. All estimates are shown over time with median (line), 50% credible intervals (dark shaded region) and 95% credible intervals (light shaded region). The grey shaded regions highlight the Autumn, Winter and Spring school holidays for each jurisdiction in 2025.

### Real-time estimates

Trends in case time series were estimated in real-time over the course of the season and included in weekly situational assessment reports provided to government partners (from 16 May-3 October 2025). The national report included: a comparison of trends in cases for each pathogen, across all eight jurisdictions, from 2023 onwards (e.g. Fig.1); and estimates of the growth rate, doubling/halving time, and probability the growth rate was greater than zero (cases increasing) on the last day of data for all pathogens and jurisdictions (e.g. Fig.4). We also estimated the proportion of influenza cases that were attributable to each influenza subtype over time.

**Figure 4.**
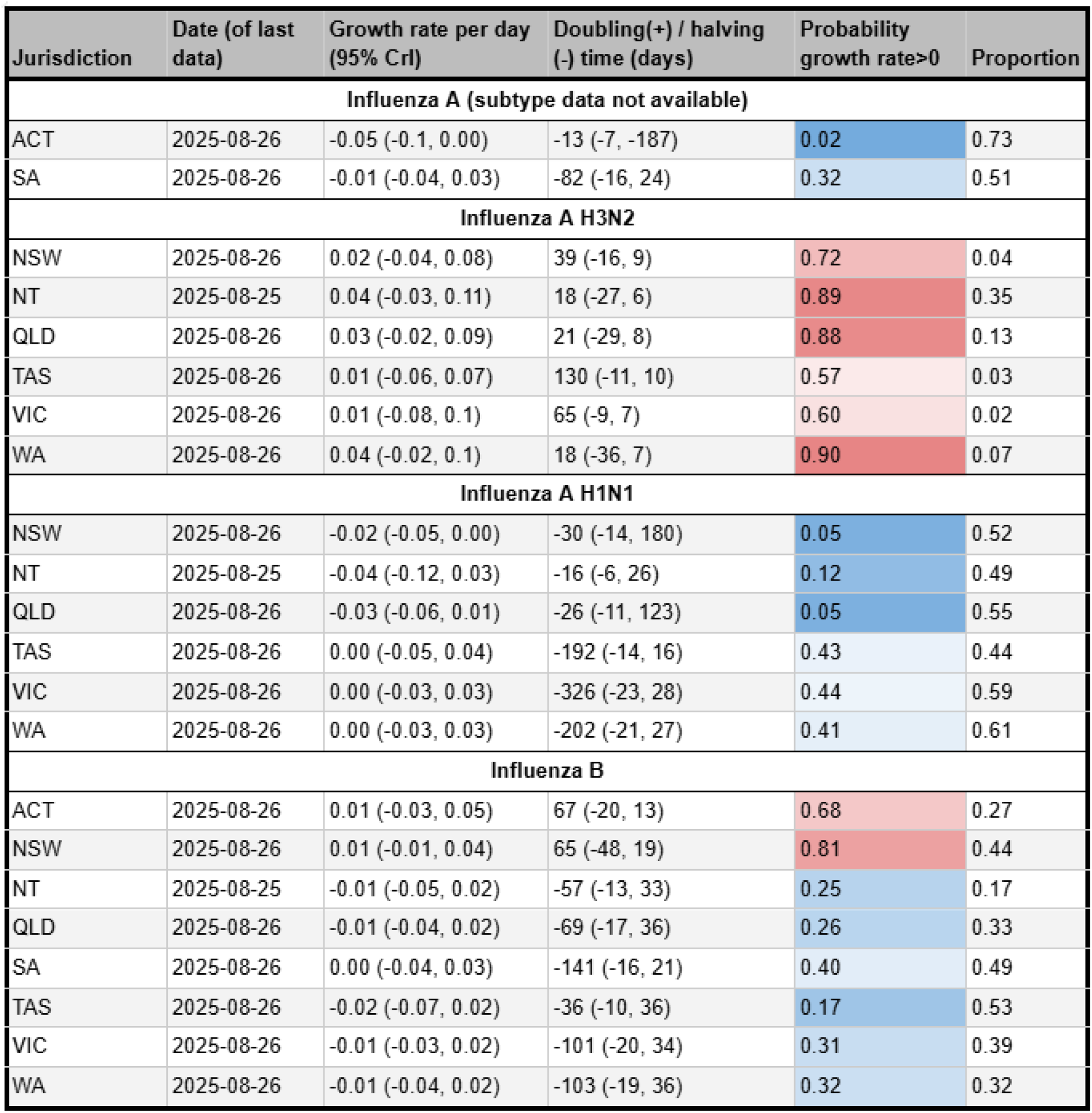
Example of tables presented in reports. An example table presented in our weekly reports including for the last day of the model fit estimates of: the growth rate, doubling/halving time, probability the growth rate was greater than zero (cases increasing) and the proportion of influenza cases that were attributable to each influenza subtype. This table was included in the report produced on 28 August 2025 (Report 16). Note we also included an additional figure detailing analogous estimates for trends in SARS-CoV-2, RSV, and overall influenza cases.

We compared real-time estimates of case trends to end of season estimates (Fig.5, SFig.15-SFig.20). We observed significant overlap in the 95% credible intervals of real-time estimates with end of season estimates for multiple jurisdictions (Fig.5 left hand panels). However, in some jurisdictions, we observed significant disagreement between real-time estimates and end of season estimates (Fig.5 right panels), likely predominantly associated with revisions to data between the data received in real-time and the end of season dataset. For example, in the six week period starting from 31 July 2025, the number of cases on the last day of data (used in the analysis) was consistently revised upwards for RSV in QLD (by 26-46 cases or 16-69%) and influenza in WA (by 53-122 cases or 46-349%); as such our real-time estimates were consistently biased downwards. For jurisdictions where minimal revisions were made to the datasets, our real-time estimates were in good agreement with end of season estimates. We also observed periods in which real-time estimates were biased due to anomalistic revisions to the data. In VIC, SARS-CoV-2 data received in real-time from 15 May-17 July (rounds 1-10) were revised downwards following routine data cleaning and re-classification from 24 July 2025 (round 11), thus our real-time estimates were often biased upwards compared to the end of season estimates. In early-July in WA, there was a single week in which SARS-CoV-2 data for the entire week were revised upwards (as opposed to just the last day) (SFig.15); this significant revision drew our attention to systematic revisions on the last day of data for SARS-CoV-2 in WA. Accordingly, for the remainder of the season we used only up to the penultimate day of data in analyses, and real-time estimates subsequently became more consistent with end of season estimates. Following retrospective analyses of the 2025 winter period, the last day of data was routinely excluded from all influenza analyses in our inter-season reports; real-time estimates in the inter-season reports were in greater agreement with end-of-season estimates compared to the within-season reports (Fig.5, SFig.19-SFig.25), except when there were anomalous data revisions (e.g. the ACT and the NT in SFig.19)

**Figure 5:**
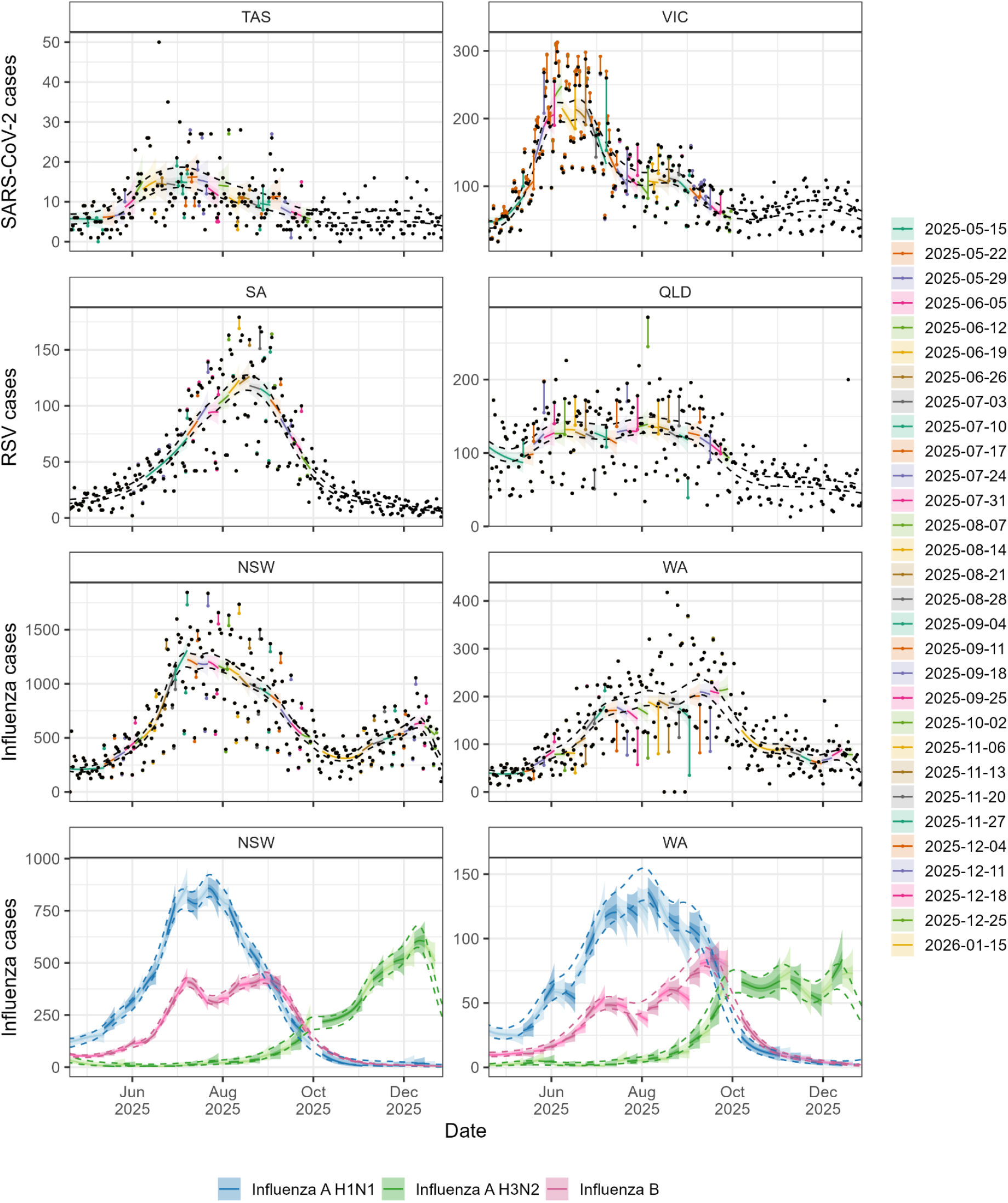
A subset of real-time estimates of case dynamics. Modelled estimates of the expected number of cases of SARS-CoV-2, RSV, influenza and influenza subtypes made in real-time (coloured lines and shaded region) for a representative subset of jurisdictions. Real-time estimates are compared to 95% credible intervals of end-of-season estimates (dashed lines). For all real-time estimates (coloured by round) we include the model posterior distribution’s median (line) and 50% and 95% credible intervals (dark shaded and light shaded regions). We include the real-time model estimates up to the final day of data used when fitting the model (for periods greater than the previous round’s model fit). The daily cases for the final end-of-season dataset (black points) and the daily cases in the real-time datasets (points coloured by round) are connected by lines to highlight revisions to data used in real-time analyses. Note that when coloured points (and connecting lines) can not be seen then there has been no revisions to the data. When a coloured point can be seen and is the same colour to the real-time estimate for the same period (and connected directly to a black data point), it indicates that the number of cases for a specific day was revised (upwards in most instances) in the following round of data, but was not revised in any further rounds of data. Note than some revisions were more complex (e.g. VIC SARS-CoV-2) and are described in more detail in Supplementary Figure 15. Real-time estimates for all jurisdictions and pathogens, and their growth rates are provided in Supplementary Figures 15-25.

Even when real-time estimates of influenza case trends were biased, real-time estimates of the relative proportions of influenza types/subtypes showed strong agreement with end of season estimates (Fig.6). In jurisdictions where real-time estimates of overall influenza case trends were consistent with end of season estimates (e.g. NSW in Fig.5), real-time estimates for influenza subtypes were also consistent with end-of-season estimates — in NSW we first estimated that influenza A H3N2 activity was most likely increasing (central estimate of growth rate above zero) in early-July (SFig.25). In jurisdictions where real-time estimates of overall influenza case trends were biased downwards relative to end of season estimates (e.g. WA in Fig.5), real-time estimates for influenza subtypes were similarly biased (SFig.21-SFig.25) (as expected). However, there was no such bias in the real-time estimates of relative proportions, with substantial overlap in the 95% credible intervals of real-time estimates and end of season estimates for most of the season. One notable exception was in QLD, where trends in influenza A H3N2 were consistently biased upwards from early-July until mid-August.

**Figure 6:**
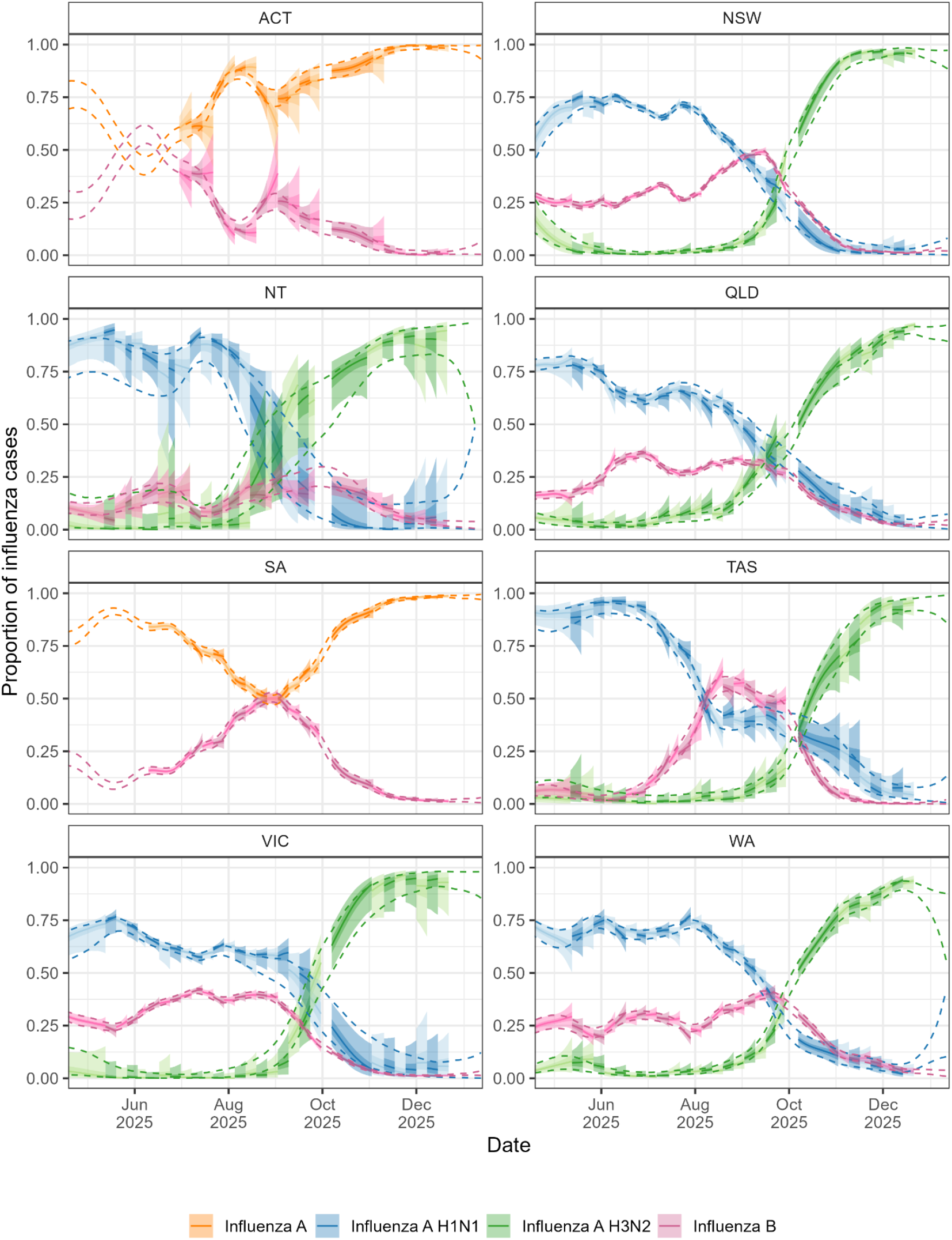
Real-time estimates of proportions of influenza subtype dynamics. Modelled estimates of the expected proportion of influenza cases attributable to each influenza subtype (coloured lines and shaded region) are compared to 95% credible intervals of end-of-season estimates (dashed lines). For all real-time estimates (alternating shades by round) we include the model posterior distribution’s median (line) and 50% and 95% credible intervals (dark shaded and light shaded regions). We include the real-time model estimates up to the final day of data used when fitting the model (for periods greater than the previous round’s model fit).

## Discussion

We have quantified the case dynamics of SARS-CoV-2, RSV, and each influenza type/subtype across all eight Australian states and territories over 2025. We employed statistical methods that robustly estimate smoothed trends (and uncertainty in those trends) while accounting for noise and day-of-the-week effects. Outputs from real-time analyses were communicated to jurisdictional and national government stakeholders via weekly reports from May to October 2025 (covering the regular respiratory virus season) and November 2025 to January 2026 (covering the period of a late influenza A H3N2 epidemic). The latter was also provided to the Australian Health Protection Committee in December 2025 to support their response to the H3N2 epidemic [14]. This reporting formed part of ACEFA’s annual Winter Situational Assessment Program—a partnership between government divisions and academic researchers to support enhanced epidemic situational awareness and response.

The case dynamics of a pathogen are the product of the underlying transmission dynamics (i.e. how infection rates change over time), and temporal trends in case-ascertainment (the proportion of infections that are identified as cases) [7,10]. As such, our analysis cannot infer the underlying infection dynamics. For some jurisdictions and pathogens, we observed highly consistent trends in case dynamics between seasons. This may reflect similarities in transmission dynamics between seasons, or similarities in case detection between seasons or various combinations (e.g. high infection rates with low case-ascertainment versus low infection rates with high case-ascertainment). Hence, identification of changes in case trends across seasons should not be interpreted as necessarily corresponding to changes in infection dynamics across seasons. Over the 2024 and 2025 seasons, multiple jurisdictions implemented RSV prevention programs including maternal vaccination and immunisation of infants at highest risk of severe RSV infection with monoclonal antibodies [15]. Our analysis cannot be used to evaluate the potential effectiveness of these programs at reducing rates of infections. Our results show comparable RSV epidemics, in terms of modelled cases, in 2023, 2024, and 2025 across all jurisdictions. Similar epidemics despite potentially modest declines in infections could be due to increasing awareness, testing, and reporting of RSV across years. Further, a decline in infections in the infant populations targeted by the RSV prevention programs may not yet be conspicuous amidst population-level case data. Additionally, overall infection levels will vary between years even in the absence of interventions — thus comparisons of infection levels between years cannot inherently disentangle the effect of an intervention from natural variation in infection levels. An epidemiological study would need to be designed and implemented to properly evaluate the effectiveness of such programs at preventing infections and hospitalisations.

Five years after its global emergence, it is still unclear if SARS-CoV-2 has settled into a regular transmission cycle in temperate Australia. Similar timings of autumn/winter (April-July) and spring/summer (October-February) epidemics were observed in the 2025, 2024 and 2023 seasons across all temperate jurisdictions; however, the increase in cases from October-December 2025 was substantially lower than in previous years. In the NT, where the majority of the population reside in tropical zones, the timing of SARS-CoV-2 epidemics was less similar between years. When repeated infections can occur in individuals, either due to the waning of immunity or emergence of new variants (or a combination), then seasonality in transmission (e.g. increased transmissibility in winter) can result in regularly timed epidemics [2,16]. The periodicity of these epidemics will depend on the strength of the seasonality and the rate at which immunity is lost or evaded [17]. A key driver of dynamics of SARS-CoV-2 cases is likely the emergence of new variants with increased transmissibility and/or immune evading properties; this season, Australian whole genome sequencing samples submitted to GISAID showed a rapid rise in the proportion belonging to lineage NB.1.8.1 [18] from April to July 2025 [19,20] — the same lineage also rose quickly to dominance in Hong Kong from February to April 2025 [19]. Post-pandemic changes to SARS-CoV-2 testing practices and test-seeking behaviour [21] is also likely a factor influencing case dynamics.

When retrospective revisions occurred to data that we received in real-time, we observed disagreement between our real-time and end of season estimates. The most common type of revision was the number of cases on the last day of full data being revised upwards in the final data set; this led to real-time estimates consistently being biased downwards in affected jurisdictions. This is likely due to delays in the processing of case notifications and incomplete recording of case received dates within all databases. This highlights the need for more systematic monitoring of differences in case numbers between datasets over the course of the season [22]. Where systematic revisions are noticed, the last day(s) of data can be routinely excluded from analyses (as done for SARS-CoV-2 in WA), preventing bias, but at the cost of less up-to-date estimates. Because our retrospective analysis of the 2025 winter period showed that the last day of data was consistently unreliable for real-time analysis, we excluded it from analyses for the inter-season influenza reports. As expected, this resulted in greater agreement between our real-time estimates and those made with the final dataset. In future seasons, statistical methods could be used to estimate a delay distribution; this would require data describing the difference between real-time datasets and future data sets; following the 2025 season we now have 31 rounds of data which can be used to parametrise such a distribution. However, these methods would only be appropriate for consistent (and therefore predictable) revisions to data in real-time, and not anomalistic revisions.

Our subtype analysis was able to provide an early signal of Australia’s unusually late and strong influenza A H3N2 epidemic. Despite limited subtyping data (e.g. 0.6% of influenza A cases were subtyped in VIC in 2025), and hence no early signal in raw case data, we first detected and reported increases in influenza A H3N2 activity in July, and by mid-August we had estimated increasing H3N2 activity across all jurisdictions where subtype data was available. This was during a period in which cases of influenza A H1N1 and B had peaked and were strongly decreasing. The combined effect of these subtype dynamics (influenza A H3N2 increasing from low levels, influenza A H1N1 and influenza B decreasing from high levels) meant that trends in overall influenza cases were observed to be decreasing. The composite indicator (overall cases) obscured the growing influenza A H3N2 epidemic for approximately two months until mid-October. This H3N2 epidemic was dominated by the novel subclade of influenza A H3N2 (‘subclade K’); subclade K was first detected in Sydney and Melbourne on 17 July 2025 and by late-October represented greater than 80% of genetically sequenced H3N2 cases in Australia [4]. Early influenza seasons, dominated by this novel subclade, were reported by northern hemisphere countries for the 2025-2026 winter season [5,6]. Increased transmissibility and/or immune evading properties of the novel subclade compared to other circulating clades/subtypes likely enabled its strong growth beyond the winter period in Australia and early season onset with high growth rates in the northern hemisphere.

This was the first season in which ACEFA reported across all eight Australian states and territories, following a pilot Situational Assessment Program in Victoria over winter 2024 [23]. As such, many data processing steps, modelling methods and reporting processes were under development during the season and areas for improvement have been identified for 2026. Real-time estimates were biased where systematic revisions to data occurred that we did not detect during the season. In future seasons we aim to develop new processes to detect revisions as soon as practicable and apply methods to account for them. We focused our 2025 analysis on the case time series for each pathogen. Other surveillance indicators for monitoring trends in respiratory pathogen activity may provide additional insights; particularly trends in pathogen specific ‘ILI+’ (i.e. the rate of influenza like illness multiplied by the test positive proportion), which has been demonstrated to be a more robust surveillance indicator for influenza-like illness causing pathogens [10]. Finally, given the known heterogeneities in infection dynamics by age, we aim to perform age-stratified analyses. This would require the development of new methodologies, including to account for contact rates within and between age groups, to jointly quantify age-stratified case dynamics.

Several of ACEFA’s government partners have highlighted the value of drawing on both within- and between-jurisdiction outputs, particularly given the synchronisation and sequencing of epidemic activity across jurisdictions. We observed that the timings of SARS-CoV-2 epidemics were similar across jurisdictions; a decline (or resurgence) of SARS-CoV-2 cases in one jurisdiction may be predictive of declines (or resurgences) in other jurisdictions. We identified transient decreases in the epidemic growth rate of influenza cases in most jurisdictions during school holidays, potentially reflecting changes in healthcare seeking behaviour or transmission (or a combination). Differential timing of school holidays occurs between jurisdictions and thus changes in dynamics (e.g. changes in growth rate) for one jurisdiction may help models predict dynamics in other jurisdictions. Over the course of the season, we identified similarities between jurisdictions in the predominant influenza subtypes; this may inform jurisdictions where subtype data are not available on the likely components of overall influenza cases. Overall, our study highlights the benefits of incorporating statistical methods into real-time surveillance of respiratory pathogen epidemics and in consolidating reporting across jurisdictions. In particular, Australia’s unusually late and strong influenza A H3N2 epidemic emphasises the importance of subtype-specific analysis for monitoring influenza activity and demonstrates the utility of our multi-subtype model [8,24] for this task.

## Funding

This research is supported by the Australian Consortium of Epidemic Forecasting and Analytics (ACEFA), a National Health and Medical Research Council of Australia Centre of Research Excellence (2035303).

OE is supported by a University of Melbourne McKenzie fellowship. FMS is supported by the National Health and Medical Research Council of Australia through the Investigator Grant Scheme (Emerging Leader Fellowship, 2021/GNT2010051). JMM is supported by the Australian Research Council through the Laureate Fellowship Scheme (FL240100126). FMS and JMM’s research is also supported by an Australian Research Council Discovery Project Grant (DP240102286). RJT was supported by a Melbourne Research Scholarship.

## Acknowledgements

National Notifiable Disease Surveillance System data were provided by the Interim Australian Centre for Disease Control, Australian Government Department of Health and Aged Care, on behalf of the Communicable Diseases Network Australia (CDNA). We thank public health staff and data managers in state and territory health departments, and the Australian Government Department of Health and Aged Care, along with state and territory public health laboratories.

Public Health Event Surveillance System (PHESS) data were provided by the Department of Health, Victoria. We further wish to acknowledge the medical practitioners and pathology services who diagnosed and notified the cases to the Department of Health, Victoria. We also acknowledge the public health officers, medical officers, epidemiologists and systems specialists who have also contributed to the collection and curation of the PHESS dataset.

Finally, we acknowledge: the Communicable Disease Control Branch, SA Health; Professor Ross Andrews of the Public Health Intelligence Branch, Population Health Division, Queensland Health; and the Preparedness, Planning and Surveillance Branch, Health and Community Services Directorate, ACT Government, particularly Rebecca Hundy, Timothy Sloan-Gardner, and Alexandra Marmor.

This research was supported by use of the Nectar Research Cloud, a collaborative Australian research platform supported by the NCRIS-funded Australian Research Data Commons (ARDC), and by the University of Melbourne Research Computing Services.

## Code and data availability

Code supporting this publication and all modelled estimates are publicly available at https://github.com/acefa-hubs/SitAssessmentPaper2025

The case dataset is not publicly available; for access to the raw data, a request must be submitted to the Australian Government Department of Health and Aged Care, which will be assessed by a data committee independent of the authorship group.

## Notes

### Competing Interest Statement

The authors have declared no competing interest.

### Author Declarations

The Human Research Ethics Committee of the University of Melbourne gave ethical approval for this work (application identifier 2024-26949-50575-3).

