## Supplementary Material for "Real-time situational assessment of respiratory virus epidemics in Australia over winter 2025 and an out-of-season influenza A H3N2 epidemic"

**Supplementary materials**

**Supplementary Tables**

| Jurisdiction | Typed (%) | Subtyped (%) |
|---|---|---|
| ACT | 90.0 | 0.0 |
| NSW | 99.7 | 2.1 |
| NT | 99.8 | 3.4 |
| QLD | 98.3 | 6.1 |
| SA | 100.0 | 0.0 |
| TAS | 100.0 | 18.4 |
| VIC | 99.9 | 0.6 |
| WA | 99.5 | 18.1 |

**Table S1: Proportion of influenza cases typed and subtyped in 2025 (1 January 2025 to 31 December 2025) to one decimal place.**

### Supplementary figures

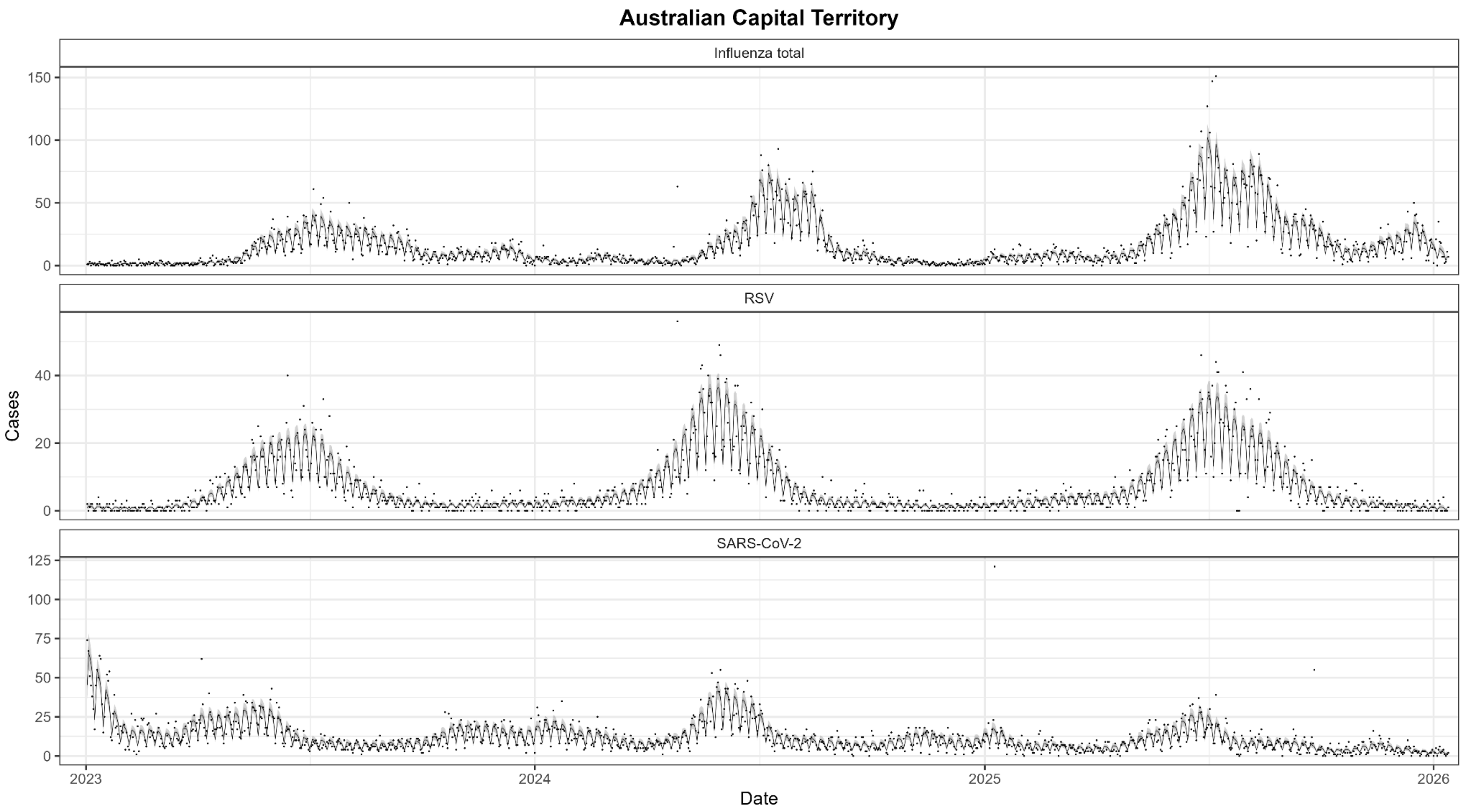


**SFig 1: Model fits to data in the Australian Capital Territory.** Modelled estimates — median estimate (line) with 95% CrI (shaded region) — of the expected number of cases over time, multiplied by estimated day of the week effects in the case data, for influenza (top), RSV (middle) and SARS-CoV-2 (bottom). Also shown are the daily number of cases by notification date (points).

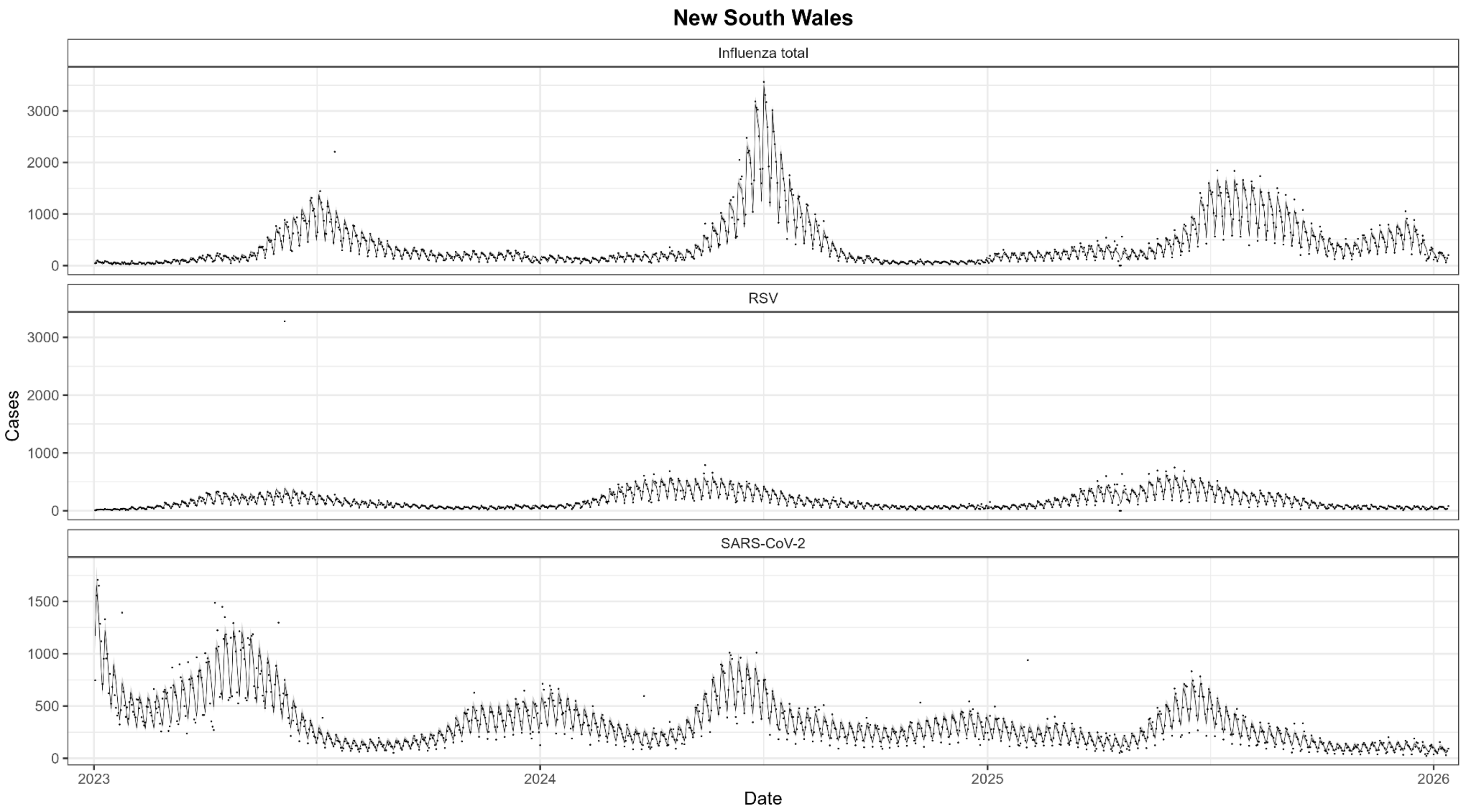


**SFig 2: Model fits to data in New South Wales.** Modelled estimates — median estimate (line) with 95% CrI (shaded region) — of the expected number of cases over time, multiplied by estimated day of the week effects in the case data, for influenza (top), RSV (middle) and SARS-CoV-2 (bottom). Also shown are the daily number of cases by notification date (points).

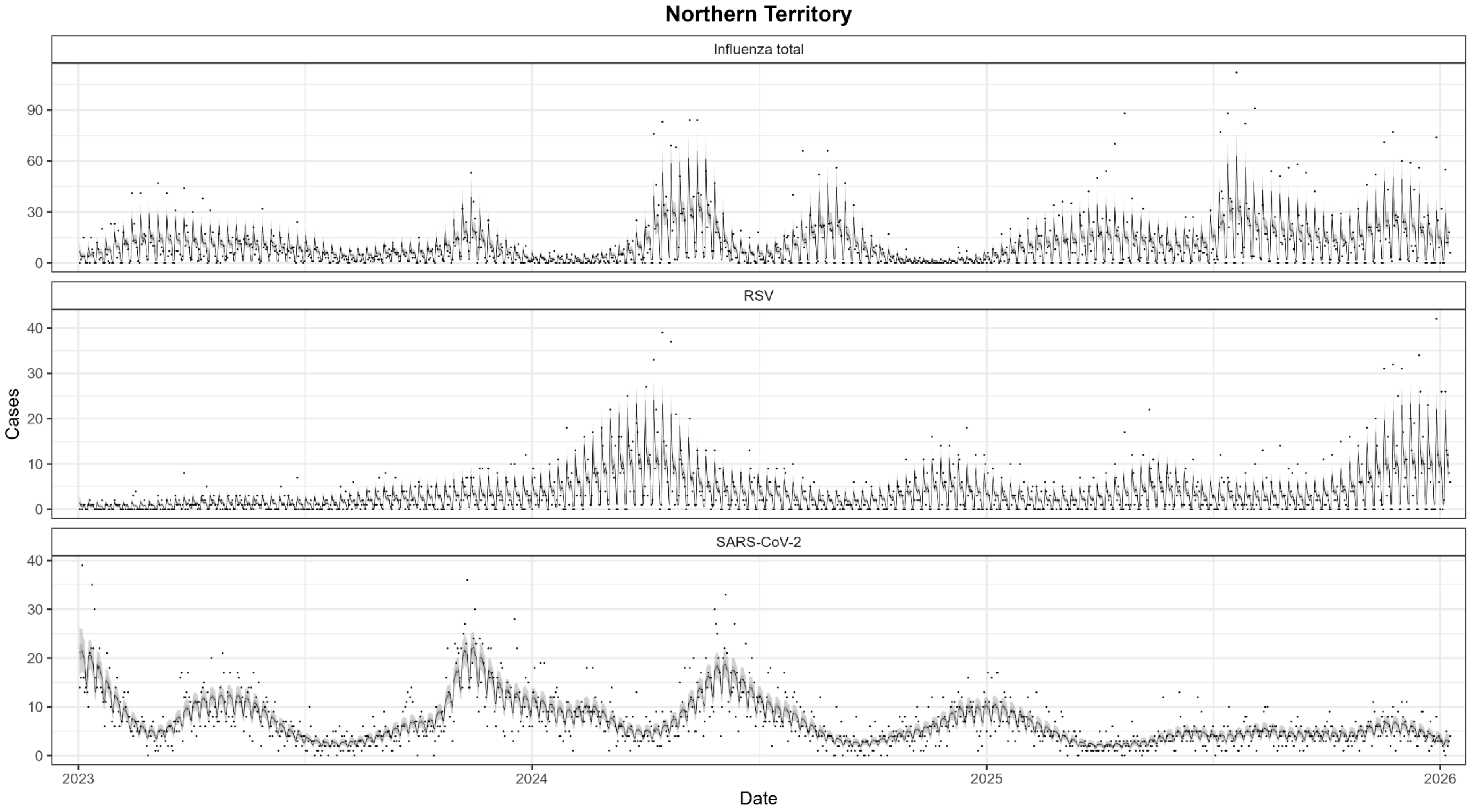


**SFig 3: Model fits to data in the Northern Territory.** Modelled estimates — median estimate (line) with 95% CrI (shaded region) — of the expected number of cases over time, multiplied by estimated day of the week effects in the case data, for influenza (top), RSV (middle) and SARS-CoV-2 (bottom). Also shown are the daily number of cases by notification date (points).

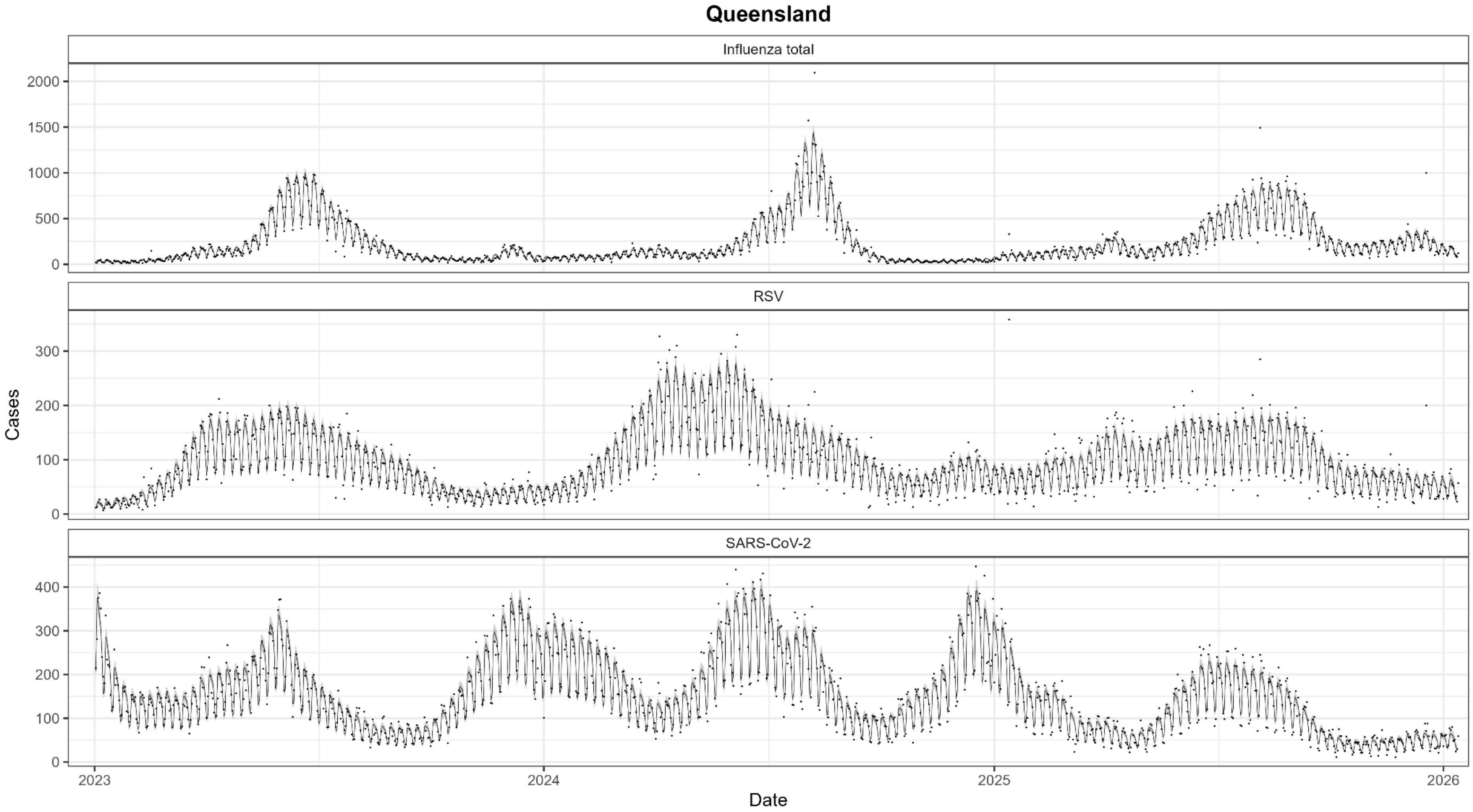


**SFig 4: Model fits to data in Queensland.** Modelled estimates — median estimate (line) with 95% CrI (shaded region) — of the expected number of cases over time, multiplied by estimated day of the week effects in the case data, for influenza (top), RSV (middle) and SARS-CoV-2 (bottom). Also shown are the daily number of cases by notification date (points).

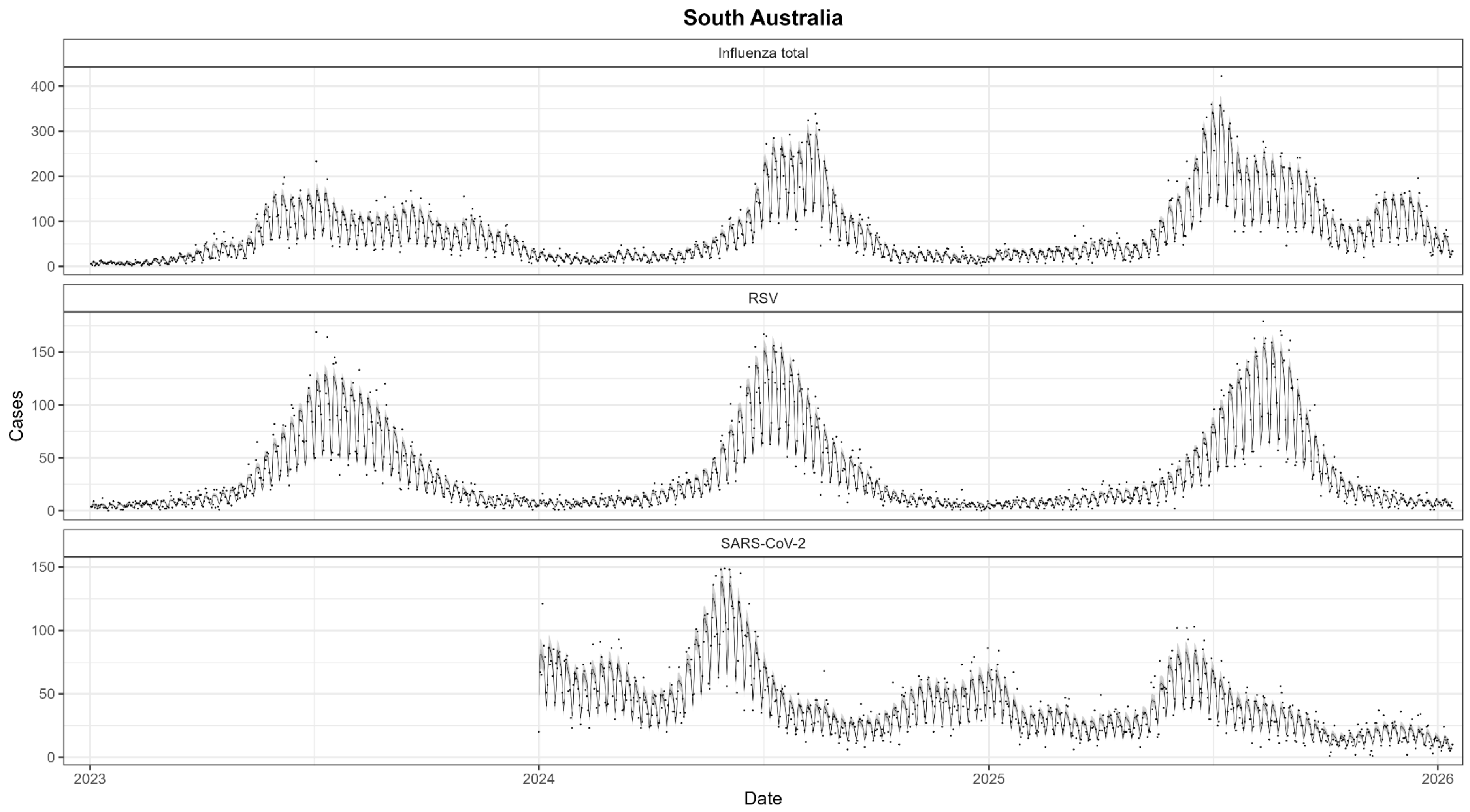


**SFig 5: Model fits to data in South Australia.** Modelled estimates — median estimate (line) with 95% CrI (shaded region) — of the expected number of cases over time, multiplied by estimated day of the week effects in the case data, for influenza (top), RSV (middle) and SARS-CoV-2 (bottom). Also shown are the daily number of cases by notification date (points).

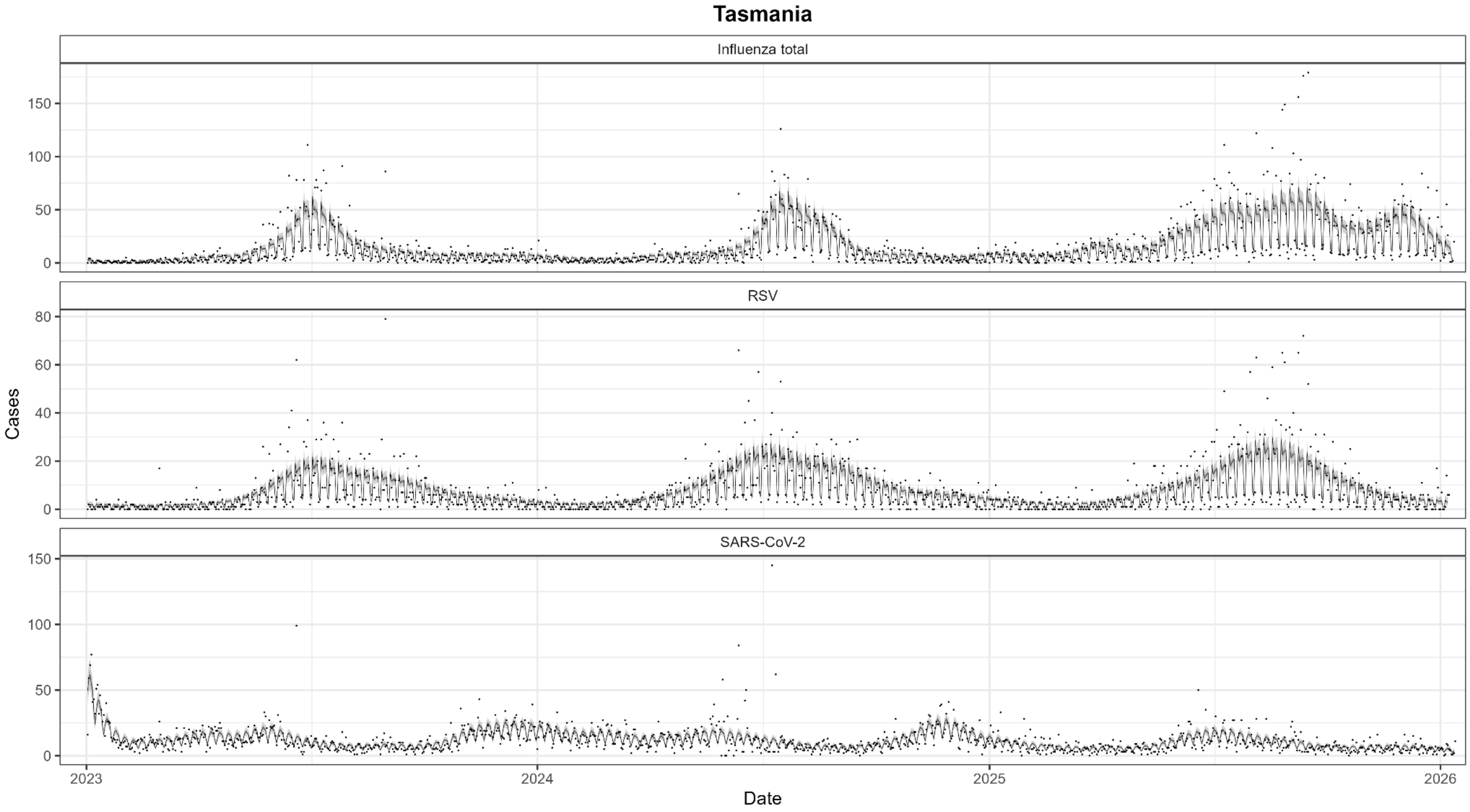


**SFig 6: Model fits to data in Tasmania.** Modelled estimates — median estimate (line) with 95% CrI (shaded region) — of the expected number of cases over time, multiplied by estimated day of the week effects in the case data, for influenza (top), RSV (middle) and SARS-CoV-2 (bottom). Also shown are the daily number of cases by notification date (points).

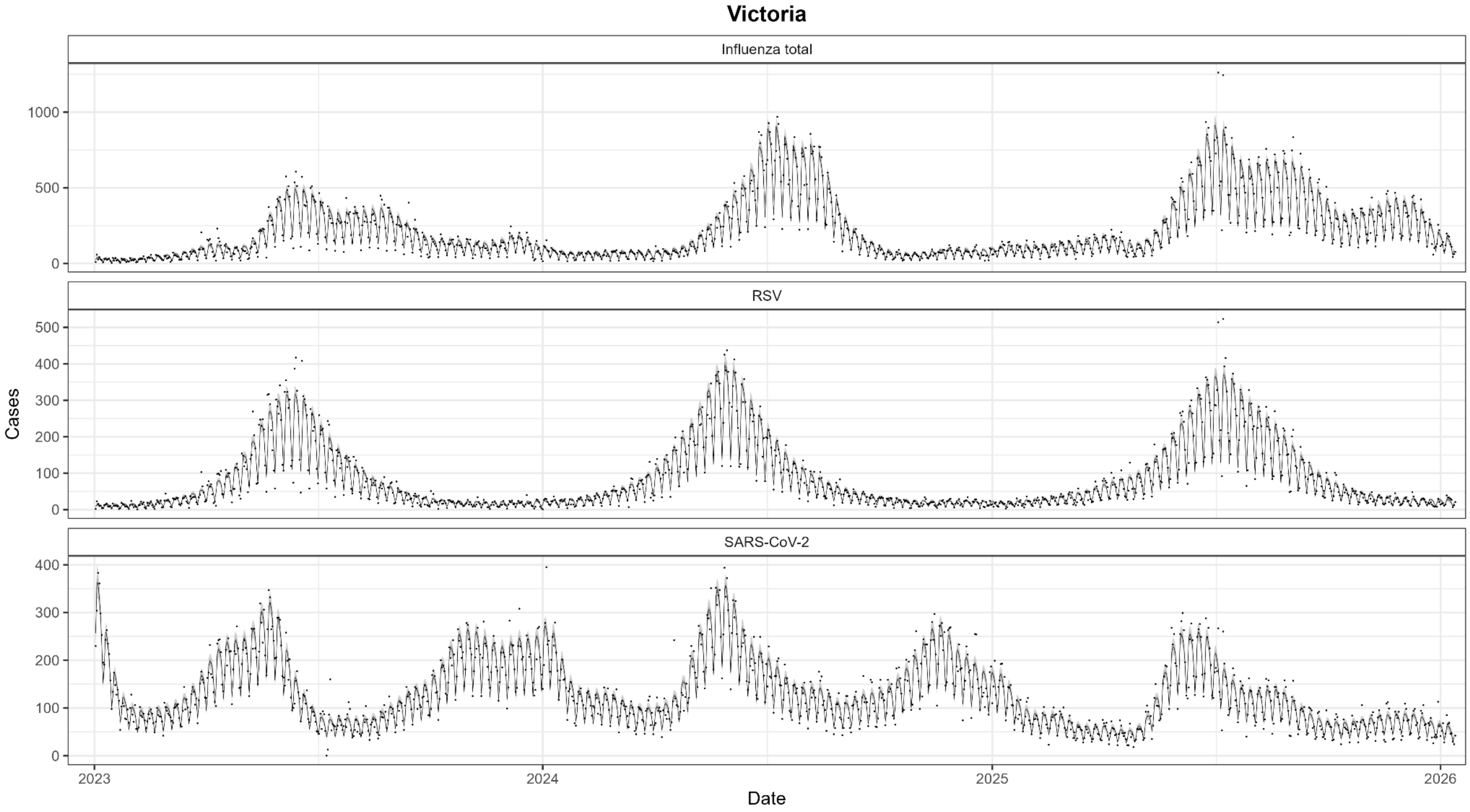


**SFig 7: Model fits to data in Victoria.** Modelled estimates — median estimate (line) with 95% CrI (shaded region) — of the expected number of cases over time, multiplied by estimated day of the week effects in the case data, for influenza (top), RSV (middle) and SARS-CoV-2 (bottom). Also shown are the daily number of cases by notification date (points).

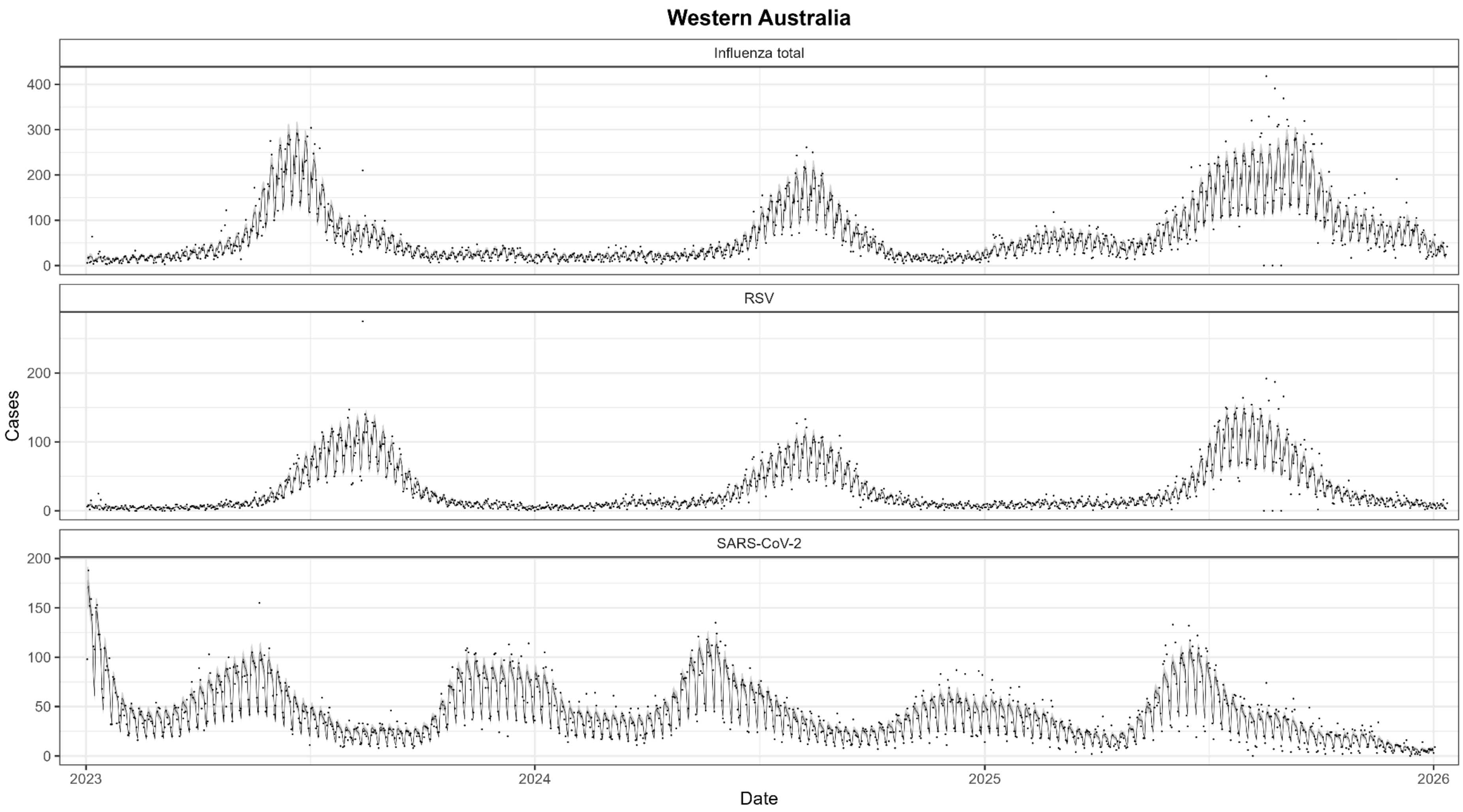


**SFig 8: Model fits to data in Western Australia.** Modelled estimates — median estimate (line) with 95% CrI (shaded region) — of the expected number of cases over time, multiplied by estimated day of the week effects in the case data, for influenza (top), RSV (middle) and SARS-CoV-2 (bottom). Also shown are the daily number of cases by notification date (points).

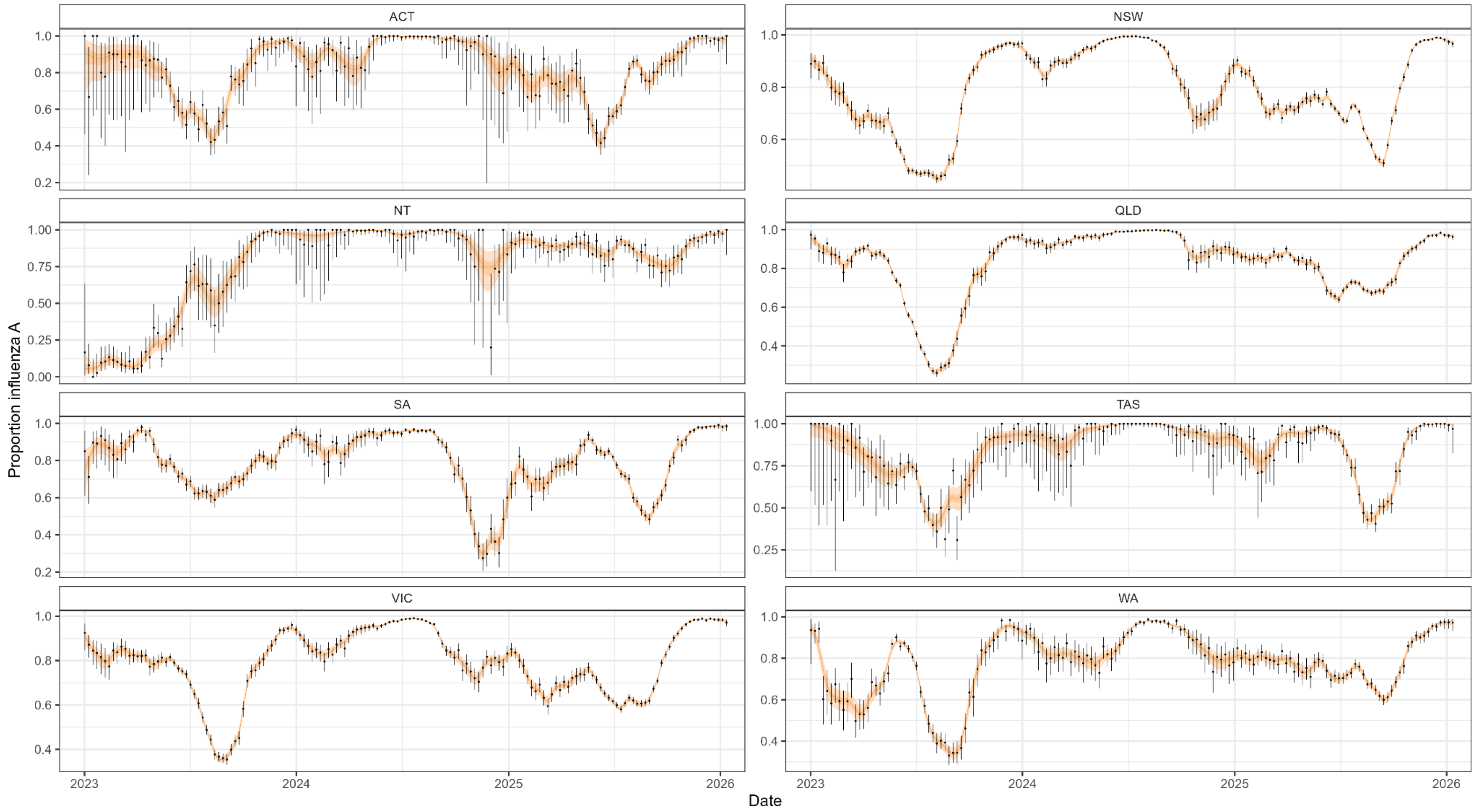


**Supplementary Figure 9: The modelled proportion of influenza attributable to influenza A.** Modelled estimates — median estimate (line) with 95% CrI (shaded region) — of the proportion of influenza cases attributable to influenza A over time for all jurisdictions. Also shown are pointwise estimates (points) and 95% binomial confidence intervals (error bars) of the weekly proportion of influenza cases attributable to influenza A in the data. Note that models were fit using daily data, but we plot weekly data which is easier to visually interpret.

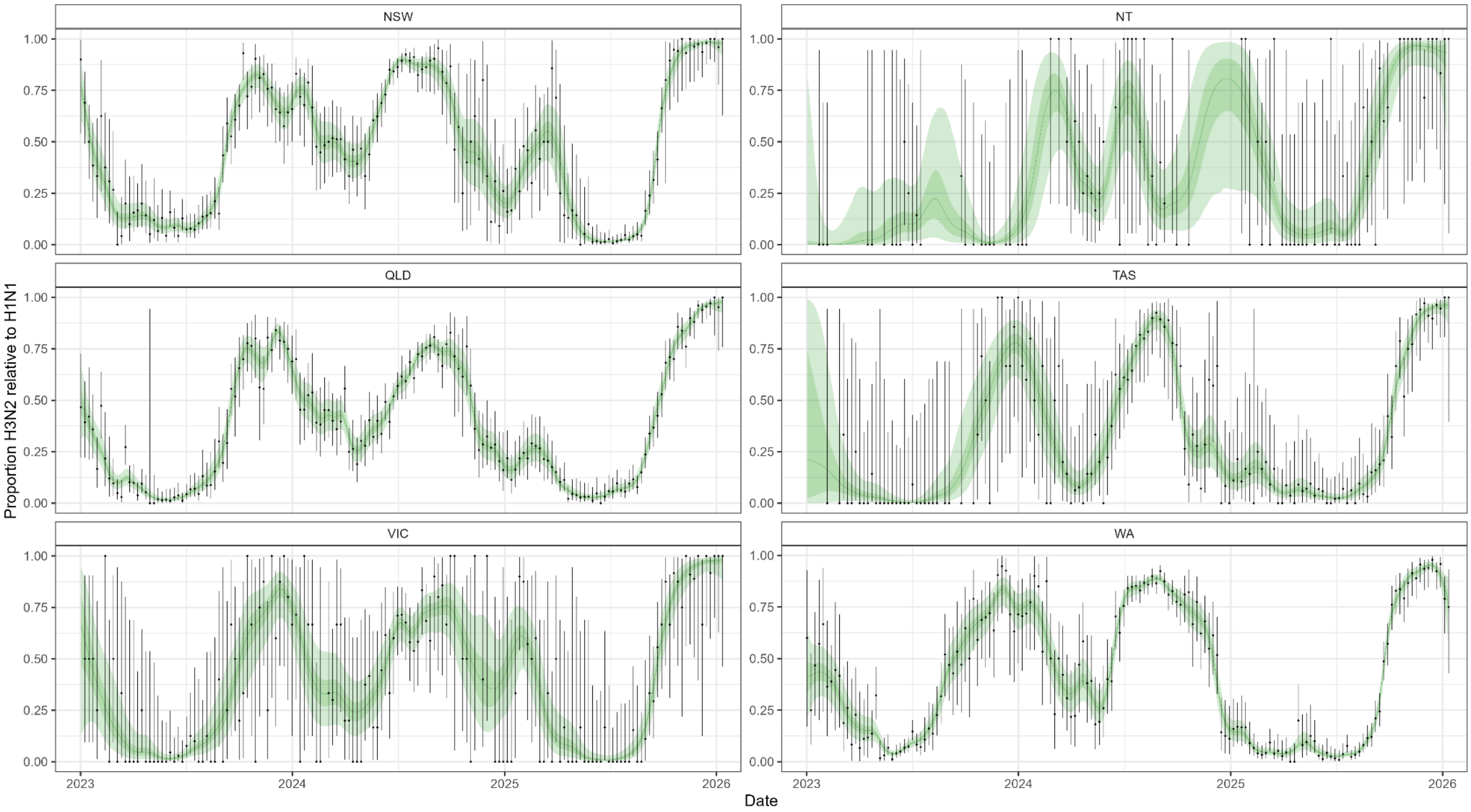


**Supplementary Figure 10: The modelled proportion of influenza A attributable to H3N2.** Modelled estimates — median estimate (line) with 95% CrI (shaded region) — of the proportion of influenza A cases attributable to H3N2 over time for all jurisdictions. Also shown are pointwise estimates (points) and 95% binomial confidence intervals (error bars) of the weekly proportion of influenza A cases attributable to H3N2 in the data. Note that models were fit using daily data, but we plot weekly data, which is easier to visually interpret.

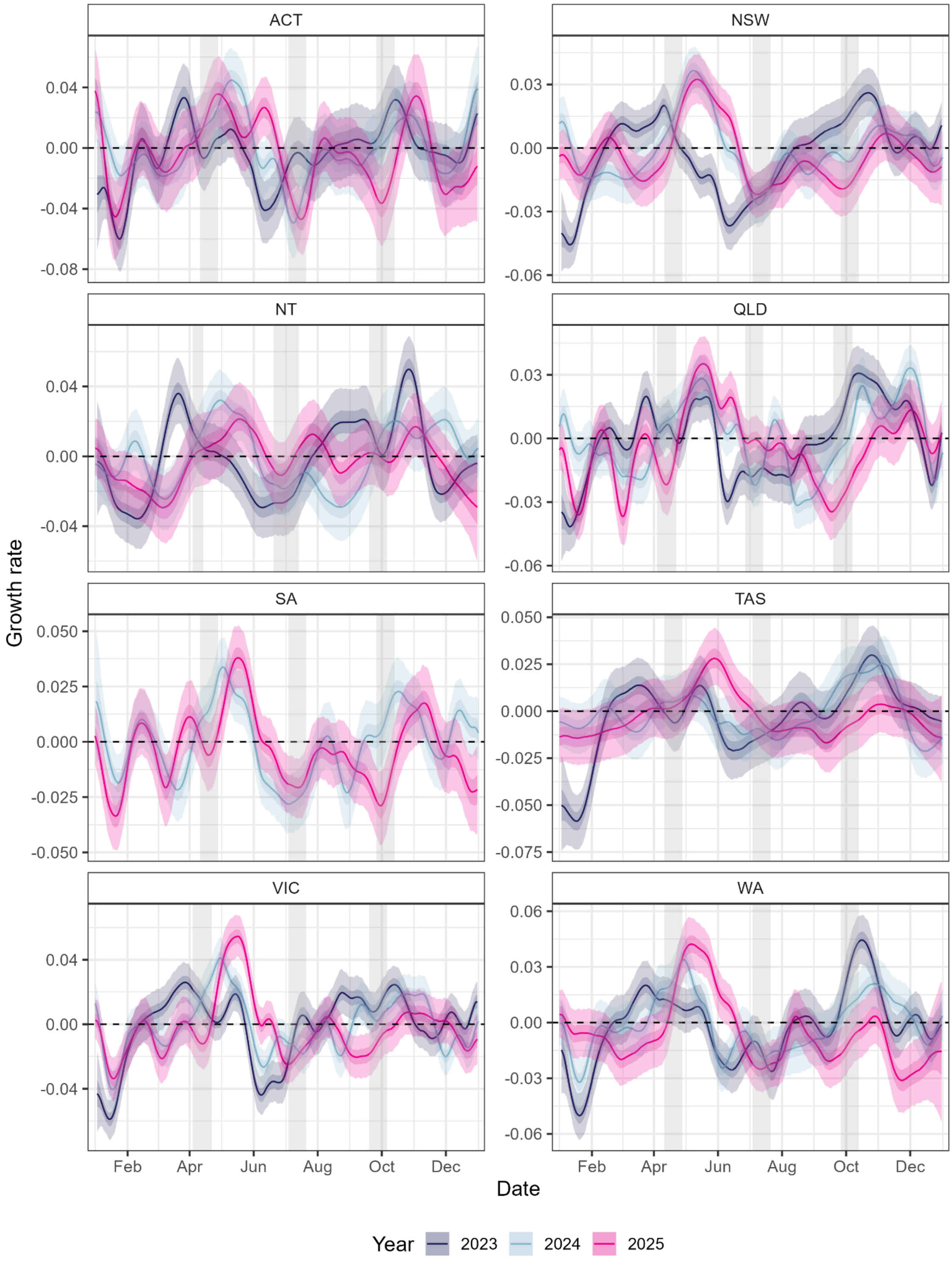


**Supplementary Figure 11: Comparison of estimated growth rates of SARS-CoV-2 over 2023, 2024 and 2025.** Modelled estimates of the growth rate of SARS-CoV-2 over time for all jurisdictions. All estimates are shown with median (line), 50% credible intervals (dark shaded region) and 95% credible intervals (light shaded region). The dashed line shows where growth rate equals 0, the transition point between epidemic growth and decline. The grey shaded regions highlight the Autumn, Winter and Spring school holidays for each jurisdiction in 2025.

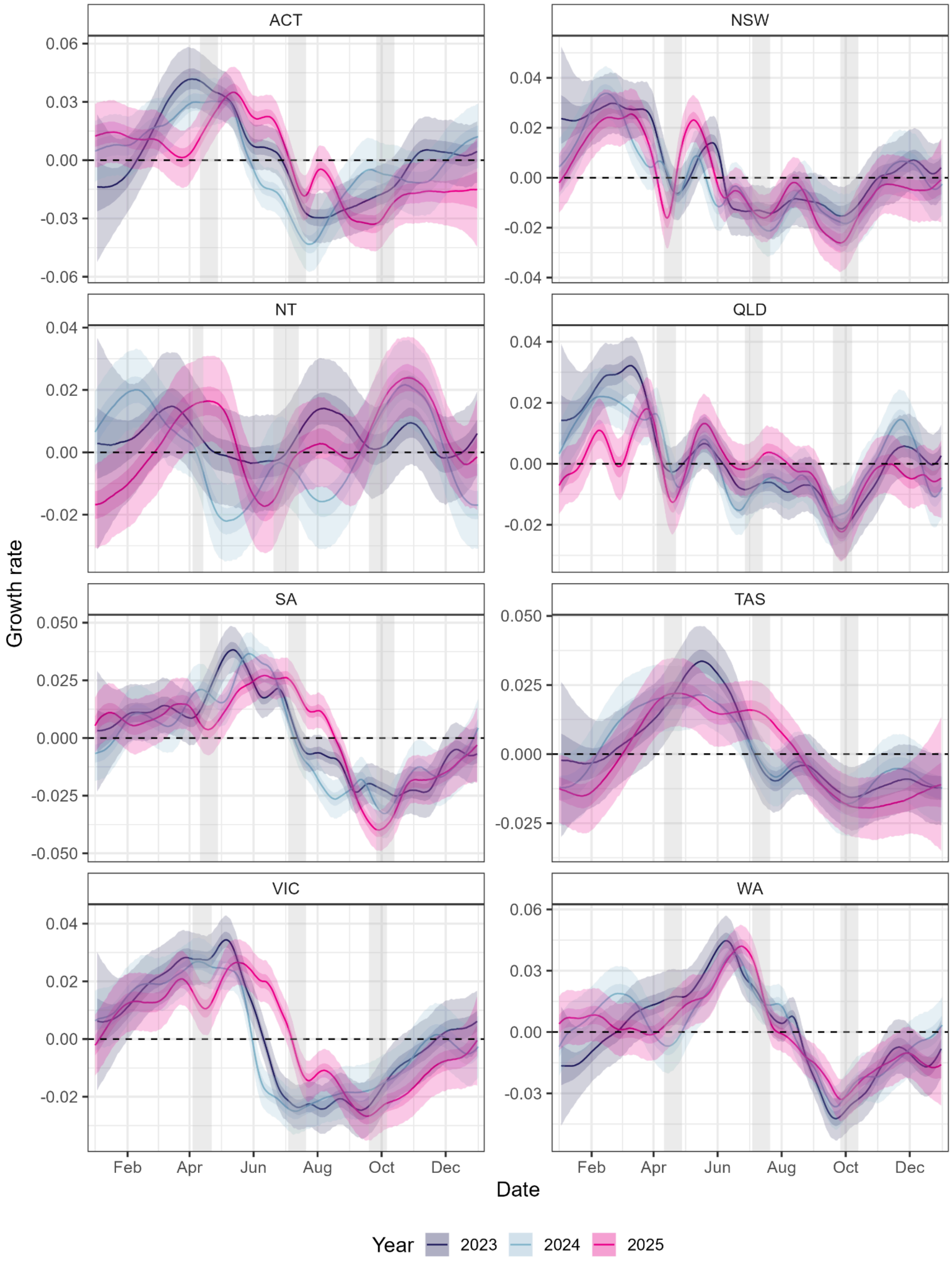


**Supplementary Figure 12: Comparison of estimated growth rates of RSV over 2023, 2024 and 2025.** Modelled estimates of the growth rate of RSV over time for all jurisdictions. All estimates are shown with median (line), 50% credible intervals (dark shaded region) and 95% credible intervals (light shaded region). The dashed line shows where growth rate equals 0, the transition point between epidemic growth and decline. The grey shaded regions highlight the Autumn, Winter and Spring school holidays for each jurisdiction in 2025.

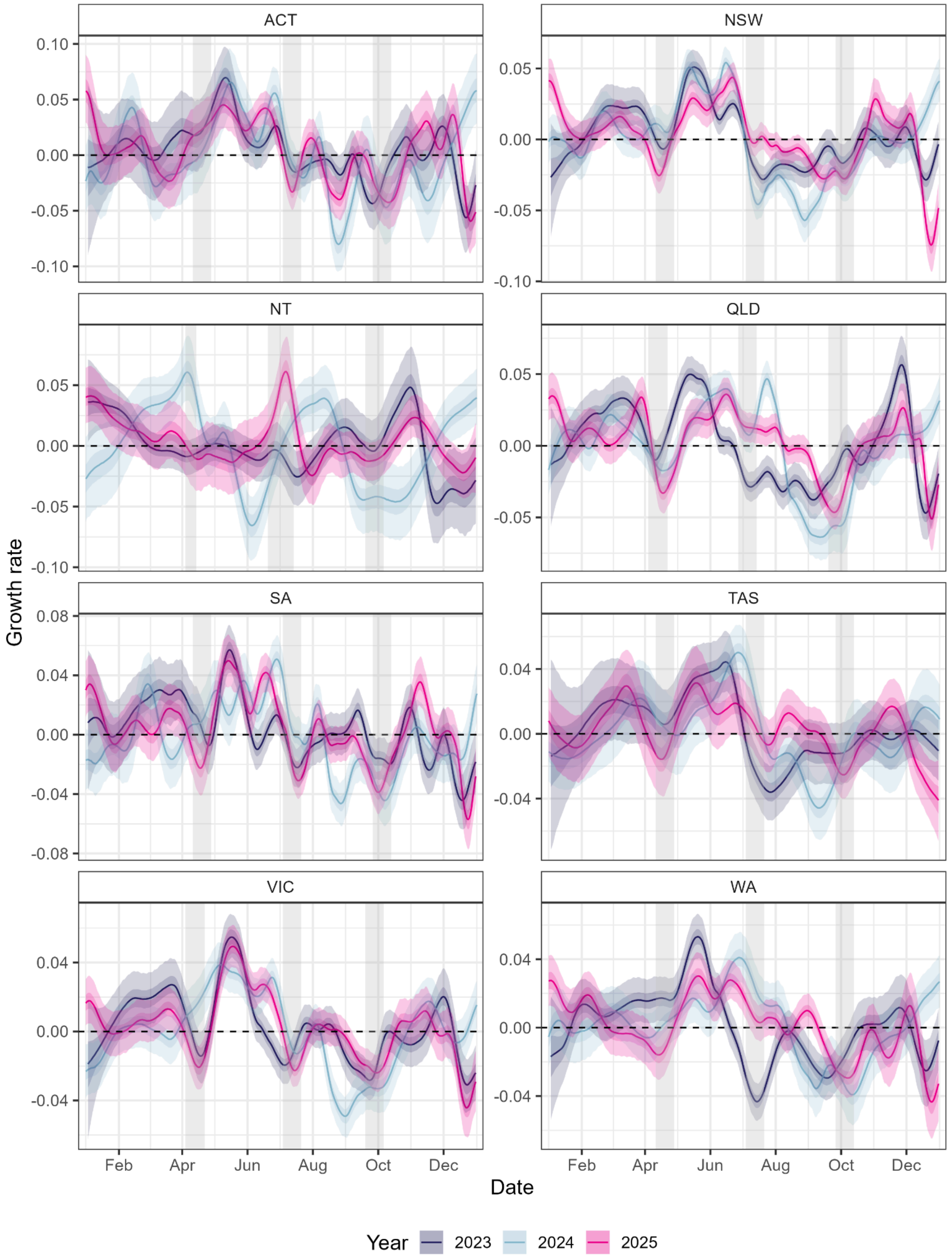


**Supplementary Figure 13: Comparison of estimated growth rates of influenza over 2023, 2024 and 2025.** Modelled estimates of the growth rate of influenza over time for all jurisdictions. All estimates are shown with median (line), 50% credible intervals (dark shaded region) and 95% credible intervals (light shaded region). The dashed line shows where growth rate equals 0, the transition point between epidemic growth and decline. The grey shaded regions highlight the Autumn, Winter and Spring school holidays for each jurisdiction in 2025.

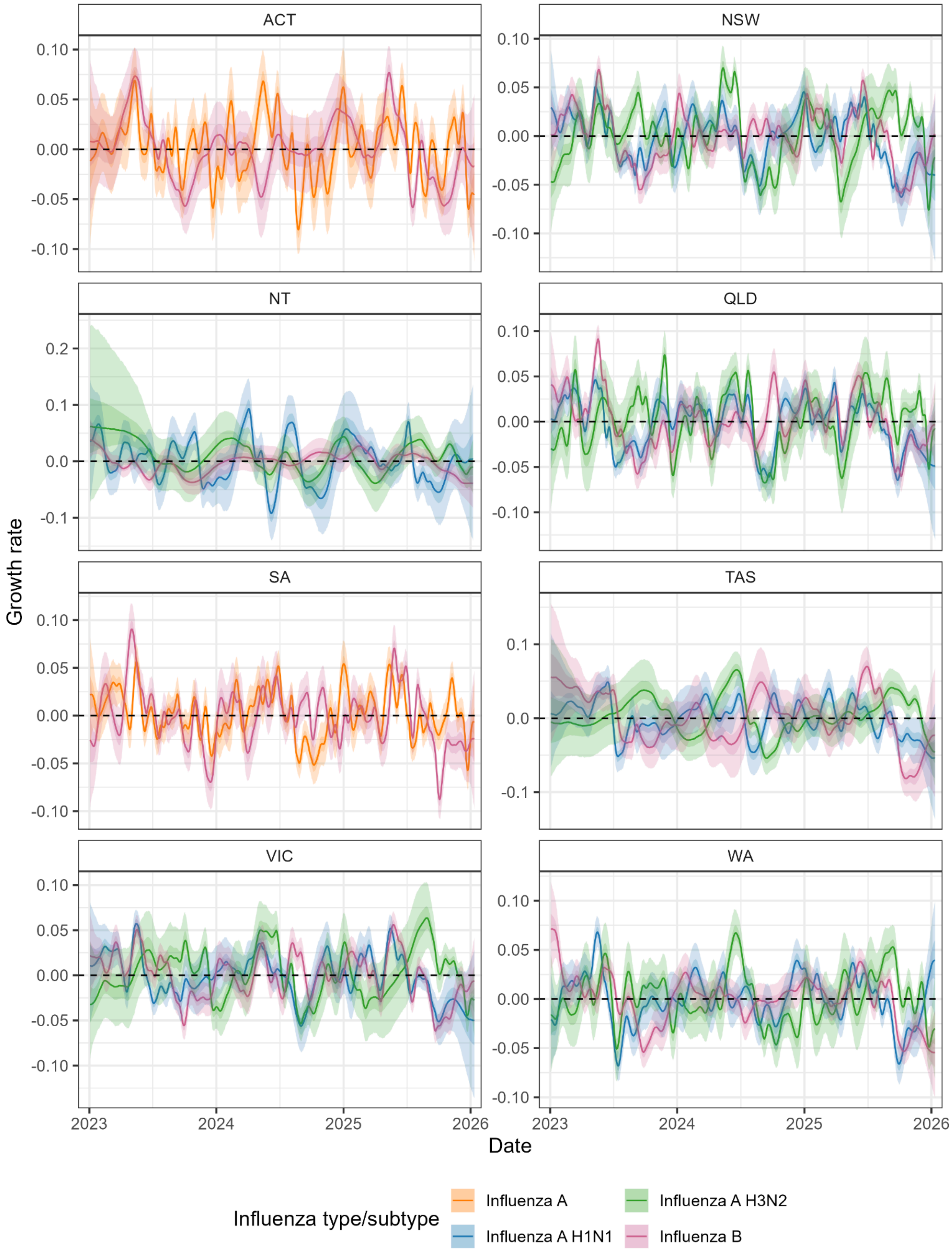


**Supplementary Figure 14: Modelled growth rate of influenza subtypes over the entire period of data.** Modelled estimates of the daily growth rate for each influenza subtype. The dashed line shows where growth rate equals 0, the transition point between epidemic growth and decline. All estimates are shown over time with median (line), 50% credible intervals (dark shaded region) and 95% credible intervals (light shaded region).

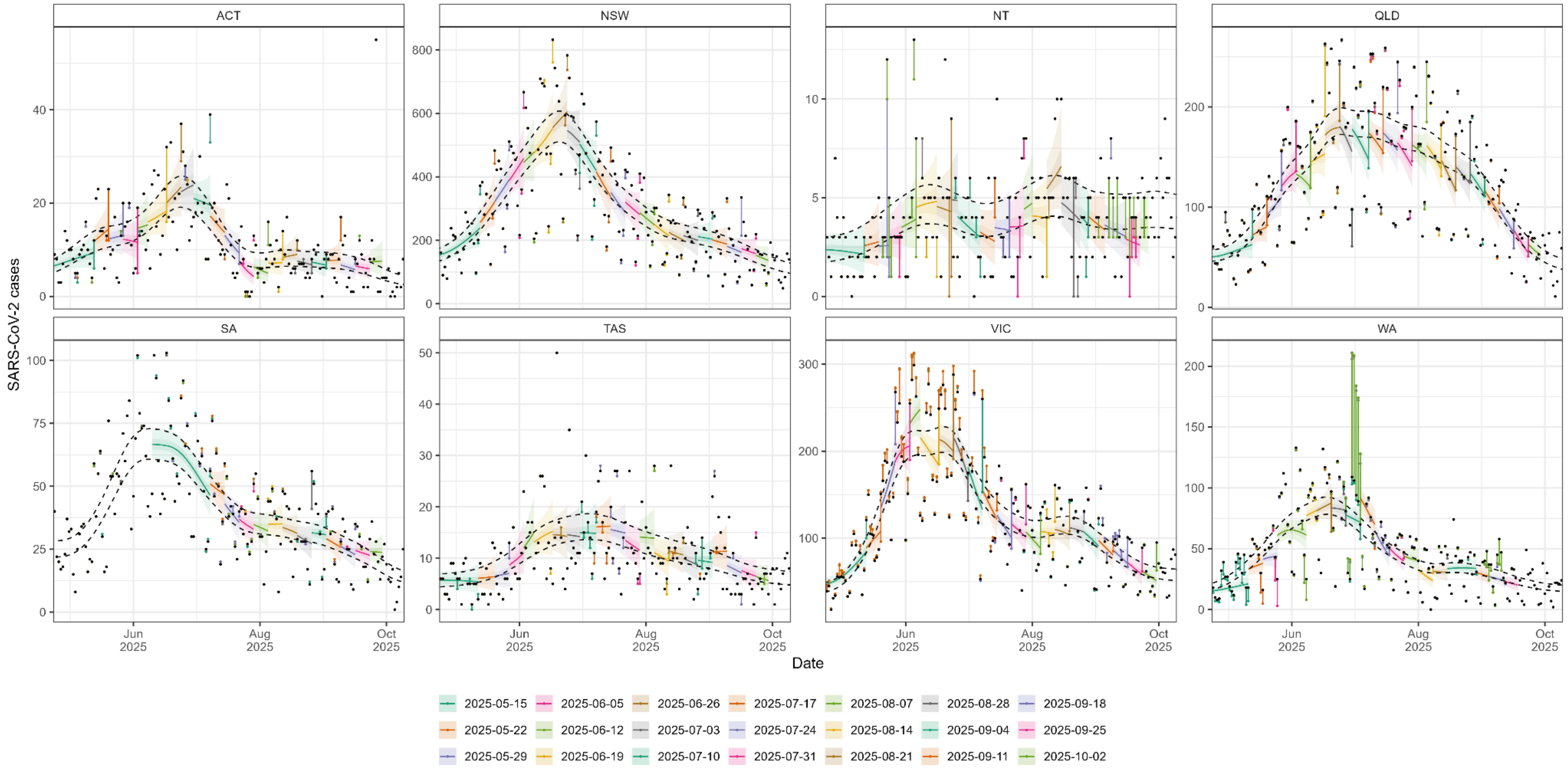


**Supplementary Figure 15: Real-time estimates of SARS-CoV-2 case dynamics.** Modelled estimates of the expected number of cases of SARS-CoV-2 made in real-time (coloured lines and shaded region) are compared to 95% credible intervals of end-of-season estimates (black dashed lines). For all real-time estimates (coloured by round) we include the model posterior distribution's median (line) and 50% and 95% credible intervals (dark shaded and light shaded regions). We include the real-time model estimates up to the final day of data used when fitting the model (for periods greater than the previous round’s model fit). The daily cases for the final end-of-season dataset (black points) and the daily cases in the real-time datasets (points coloured by round) are connected by lines to highlight revisions to data used in real-time analyses.

Note that when coloured points (and connecting lines) can not be seen then there has been no revisions to the data. When a coloured point can be seen and is the same colour to the real-time estimate for the same period (and connected directly to a black data point), it indicates that the number of cases for a specific day was revised (upwards in most instances) in the following round of data, but was not revised in any further rounds of data. There are some examples of where data was revised in multiple rounds. For example: in VIC data for multiple rounds were revised downwards in data received on 24 July 2025 (round 11) and then not revised further in future rounds; and in WA the last week of data received on 10 July 2025 (round 9) was revised significantly upwards from 17 July (round 10), before being revised significantly downwards in the final data received (note revision could have occurred in NNDSS between final round of real-time analysis for SARS-CoV-2, 2 October 2025, and the date final data was received, 14 January 2026).

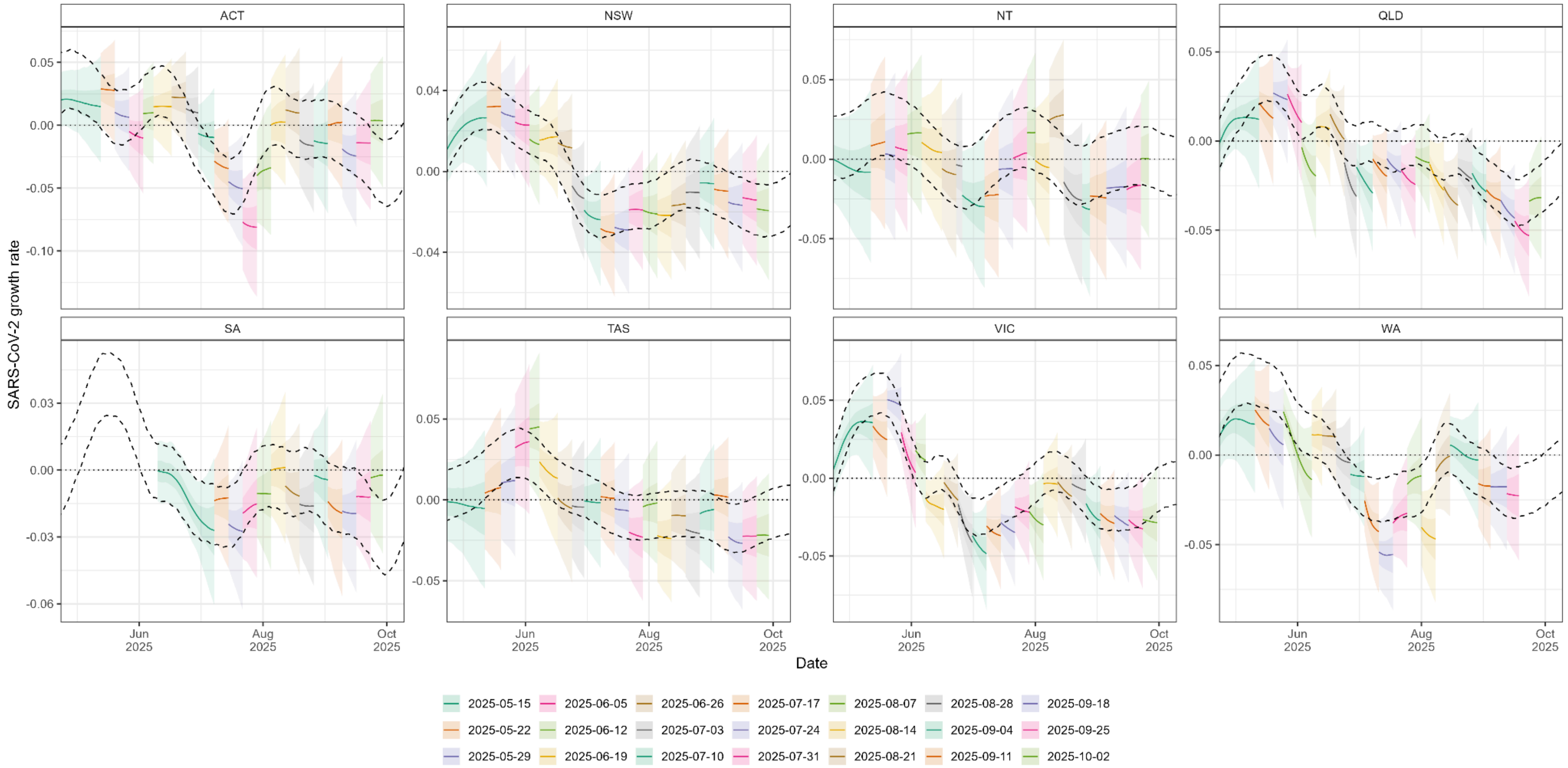


**Supplementary Figure 16: Real-time estimates of the growth rate of SARS-CoV-2 cases.** Growth rate (lines and shaded region) inferred from modelled trends in SARS-CoV-2 cases for all jurisdictions. Real-time estimates (coloured lines and shaded regions) are compared to 95% credible intervals of end-of-season estimates (black dashed lines). For all real-time estimates (coloured by round) we include the model posterior distribution's median (line) and 50% and 95% credible intervals (dark shaded and light shaded regions). We include the real-time model estimates up to the final day of data used when fitting the model (for periods greater than the previous round’s model fit).

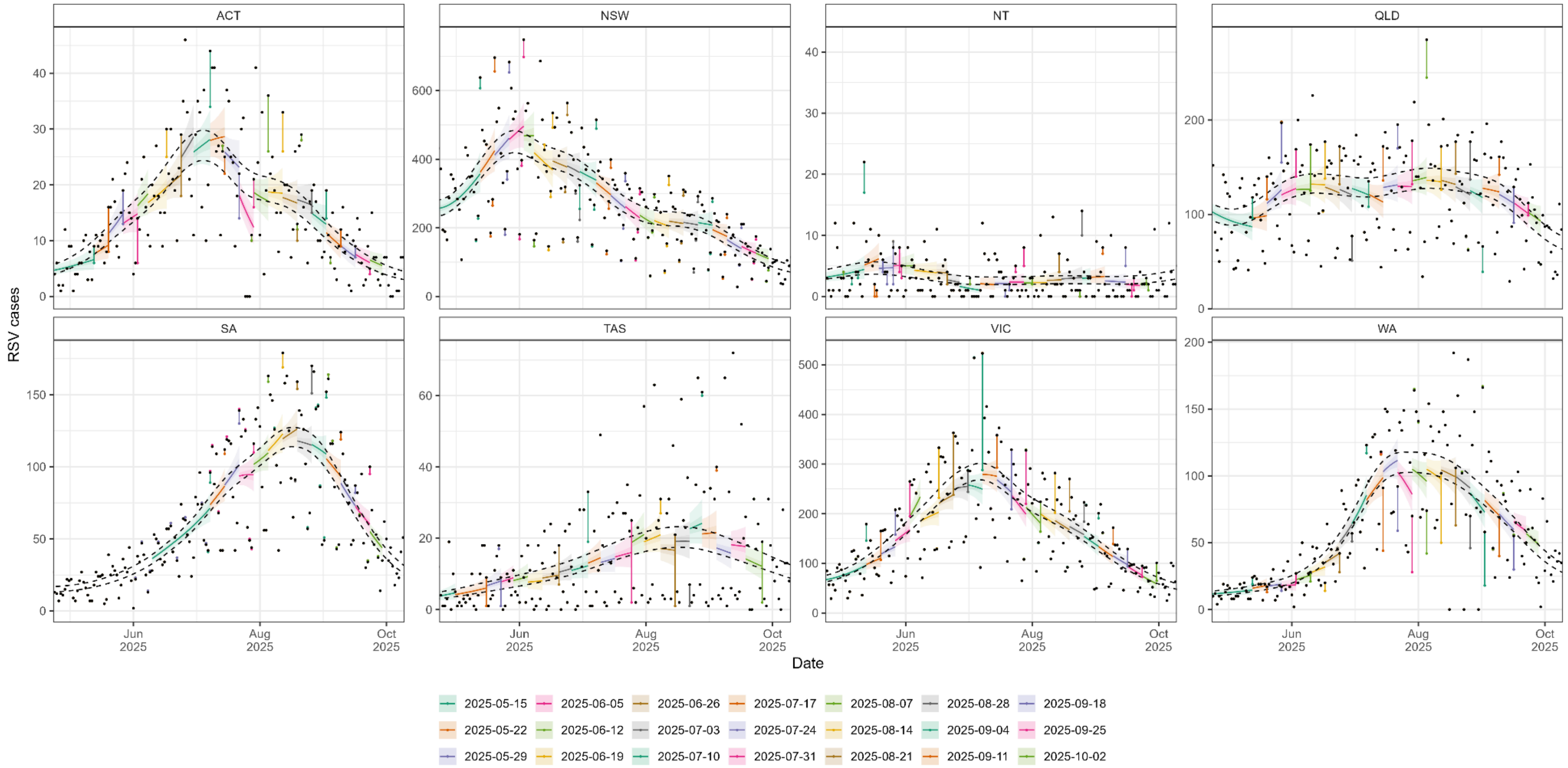


**Supplementary Figure 17: Real-time estimates of RSV case dynamics.** Modelled estimates of the expected number of cases of RSV made in real-time (coloured lines and shaded region) are compared to 95% credible intervals of end-of-season estimates (black dashed lines). For all real-time estimates (coloured by round) we include the model posterior distribution's median (line) and 50% and 95% credible intervals (dark shaded and light shaded regions). We include the real-time model estimates up to the final day of data used when fitting the model (for periods greater than the previous round's model fit). The daily cases for the final end-of-season dataset (black points) and the daily cases in the real-time datasets (points coloured by round) are connected by lines to highlight revisions to data used in real-time analyses. Note that when coloured points (and connecting lines) can not be seen then there has been no revisions to the data. When a coloured point can be seen and is the same colour to the real-time estimate for the same period (and connected directly to a black data point), it indicates that the number of cases for a specific day was revised (upwards in most instances) in the following round of data, but was not revised in any further rounds of data.

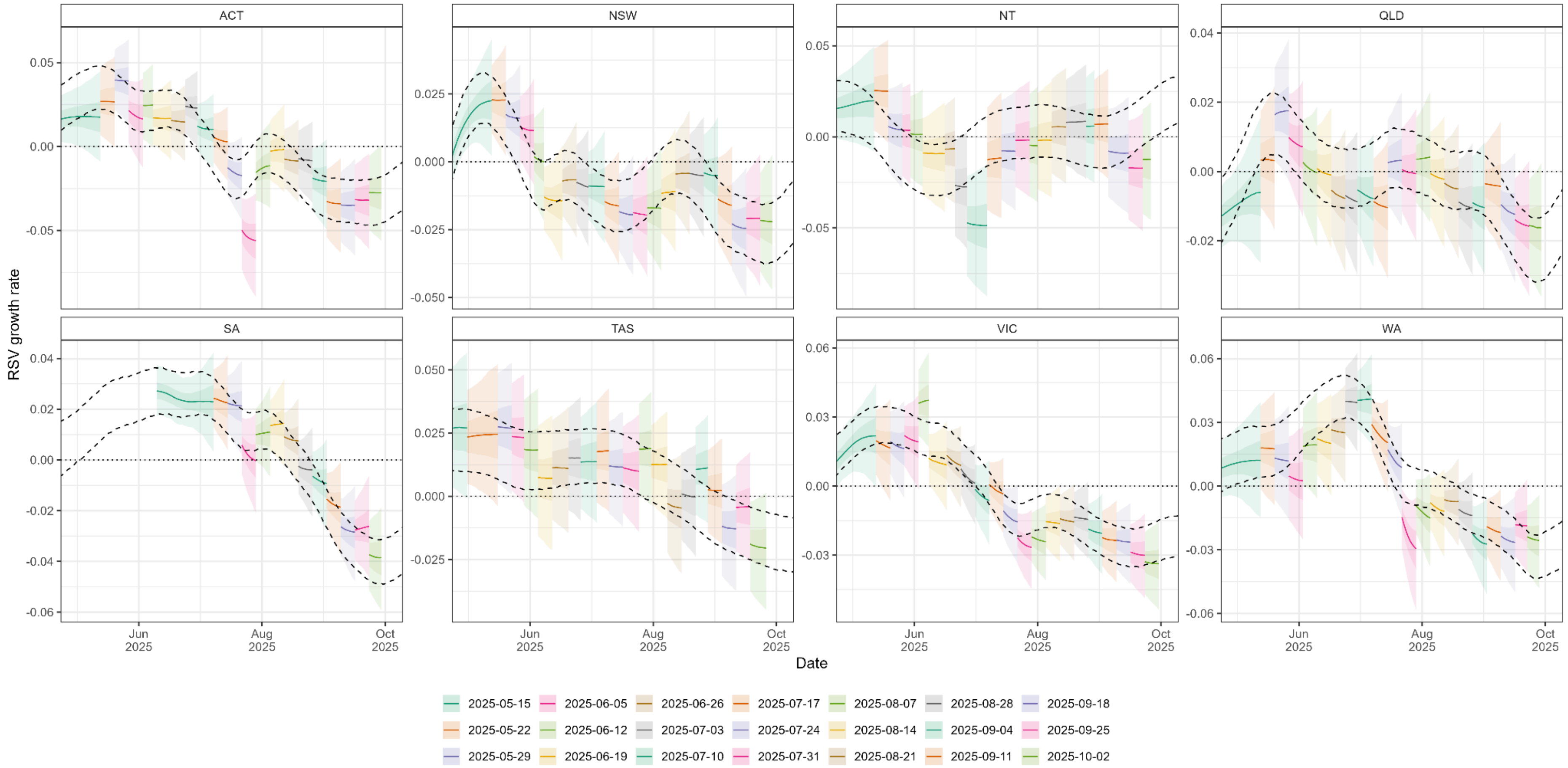


**Supplementary Figure 18: Real-time estimates of the growth rate of RSV cases.** Growth rate (lines and shaded region) inferred from modelled trends in RSV cases for all jurisdictions. Real-time estimates (coloured lines and shaded regions) are compared to 95% credible intervals of end-of-season estimates (black dashed lines). For all real-time estimates (coloured by round) we include the model posterior distribution's median (line) and 50% and 95% credible intervals (dark shaded and light shaded regions). We include the real-time model estimates up to the final day of data used when fitting the model (for periods greater than the previous round's model fit).

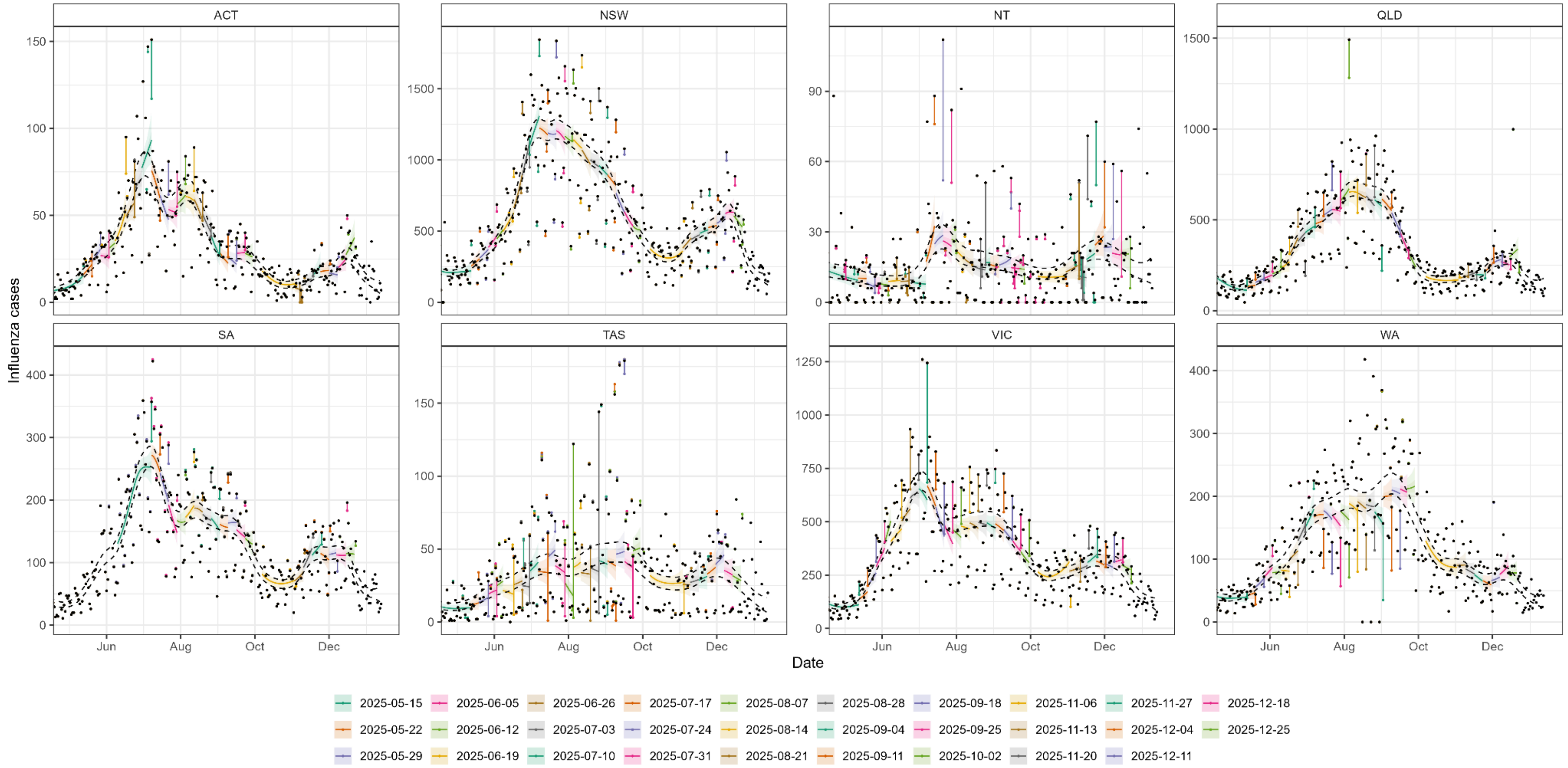


**Supplementary Figure 19: Real-time estimates of overall influenza case dynamics.** Modelled estimates of the expected number of cases of influenza made in real-time (coloured lines and shaded region) are compared to 95% credible intervals of end-of-season estimates (black dashed lines). For all real-time estimates (coloured by round) we include the model posterior distribution's median (line) and 50% and 95% credible intervals (dark shaded and light shaded regions). We include the real-time model estimates up to the final day of data used when fitting the model (for periods greater than the previous round’s model fit). The daily cases for the final end-of-season dataset (black points) and the daily cases in the real-time datasets (points coloured by round) are connected by lines to highlight revisions to data used in real-time analyses. Note that when coloured points (and connecting lines) can not be seen then there has been no revisions to the data. When a coloured point can be seen and is the same colour to the real-time estimate for the same period (and connected directly to a black data point), it indicates that the number of cases for a specific day was revised (upwards in most instances) in the following round of data, but was not revised in any further rounds of data.

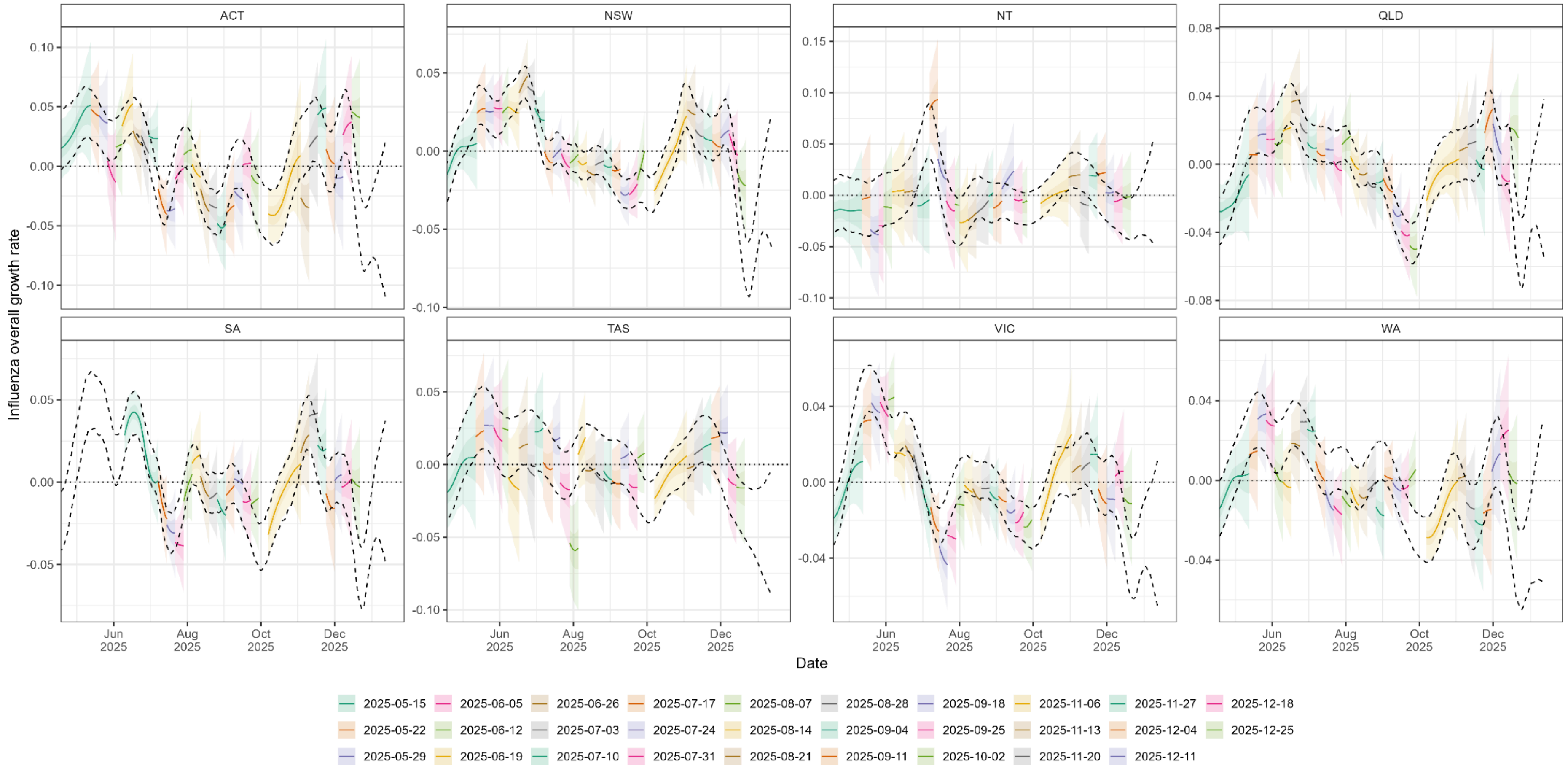


**Supplementary Figure 20: Real-time estimates of the growth rate of overall influenza cases.** Growth rate (lines and shaded region) inferred from modelled trends in overall influenza cases for all jurisdictions. Real-time estimates (coloured lines and shaded regions) are compared to 95% credible intervals of end-of-season estimates (black dashed lines). For all real-time estimates (coloured by round) we include the model posterior distribution's median (line) and 50% and 95% credible intervals (dark shaded and light shaded regions). We include the real-time model estimates up to the final day of data used when fitting the model (for periods greater than the previous round's model fit).

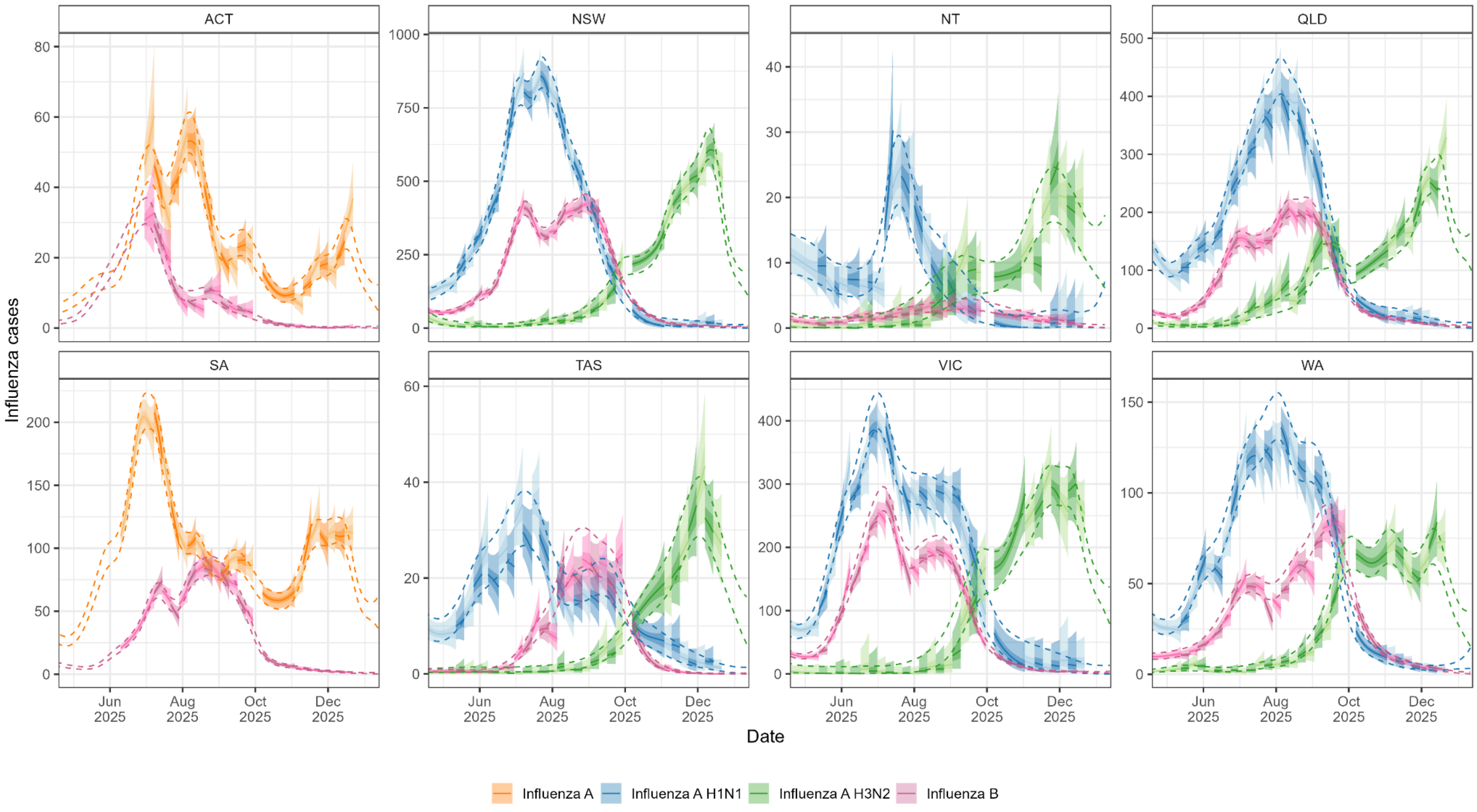


**Supplementary Figure 21: Real-time estimates of influenza subtype case dynamics.** Modelled estimates of the expected number of cases (coloured lines and shaded region) over time for each influenza subtype (colour) are compared to 95% credible intervals of end-of-season estimates (dashed lines). For all real-time estimates (alternating shades by round) we include the model posterior distribution's median (line) and 50% and 95% credible intervals (dark shaded and light shaded regions). We include the real-time model estimates up to the final day of data used when fitting the model (for periods greater than the previous round's model fit).

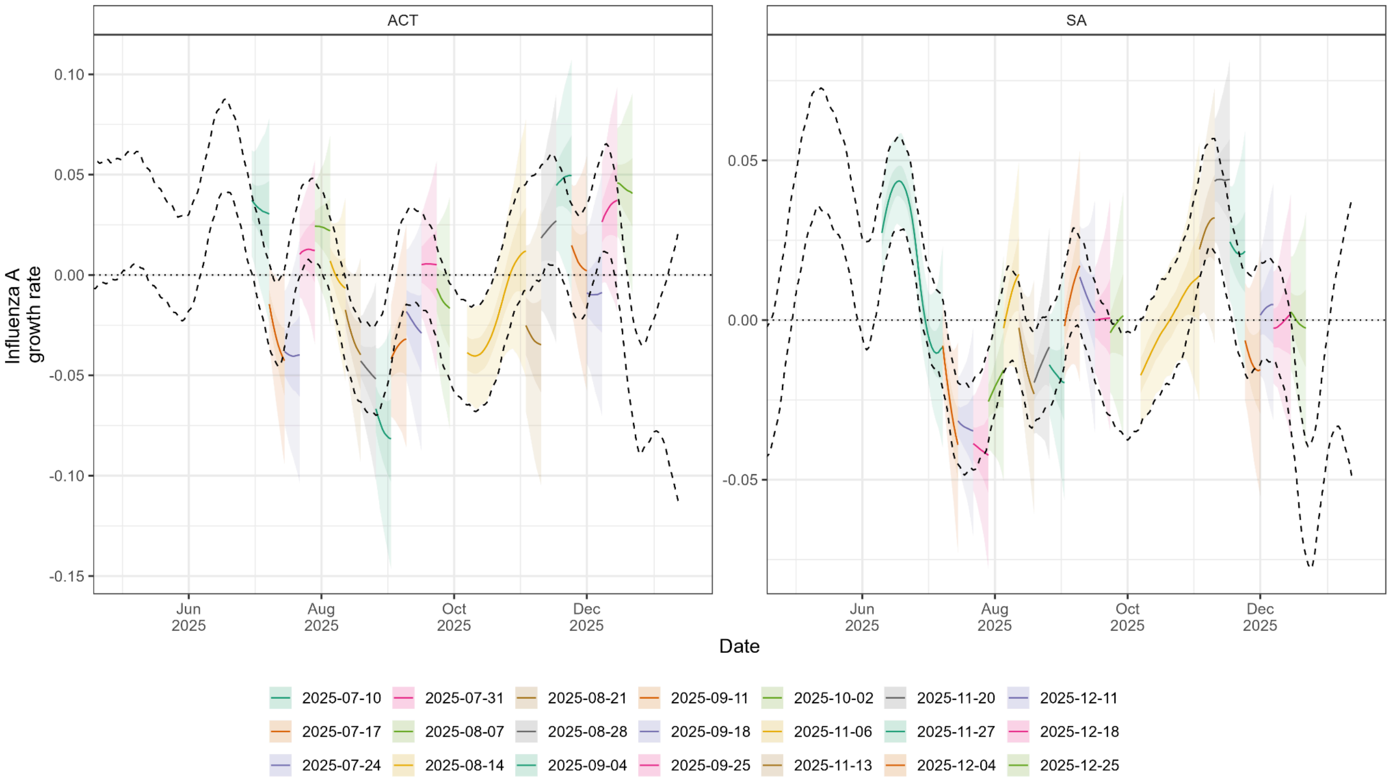


**Supplementary Figure 22: Real-time estimates of the growth rate of influenza A where subtype data is not available.** Growth rate (lines and shaded region) inferred from modelled trends in influenza A cases for all jurisdictions where subtype data were not available. Real-time estimates (coloured lines and shaded regions) are compared to 95% credible intervals of end-of-season estimates (black dashed lines). For all real-time estimates (coloured by round) we include the model posterior distribution's median (line) and 50% and 95% credible intervals (dark shaded and light shaded regions). We include the real-time model estimates up to the final day of data used when fitting the model (for periods greater than the previous round's model fit).

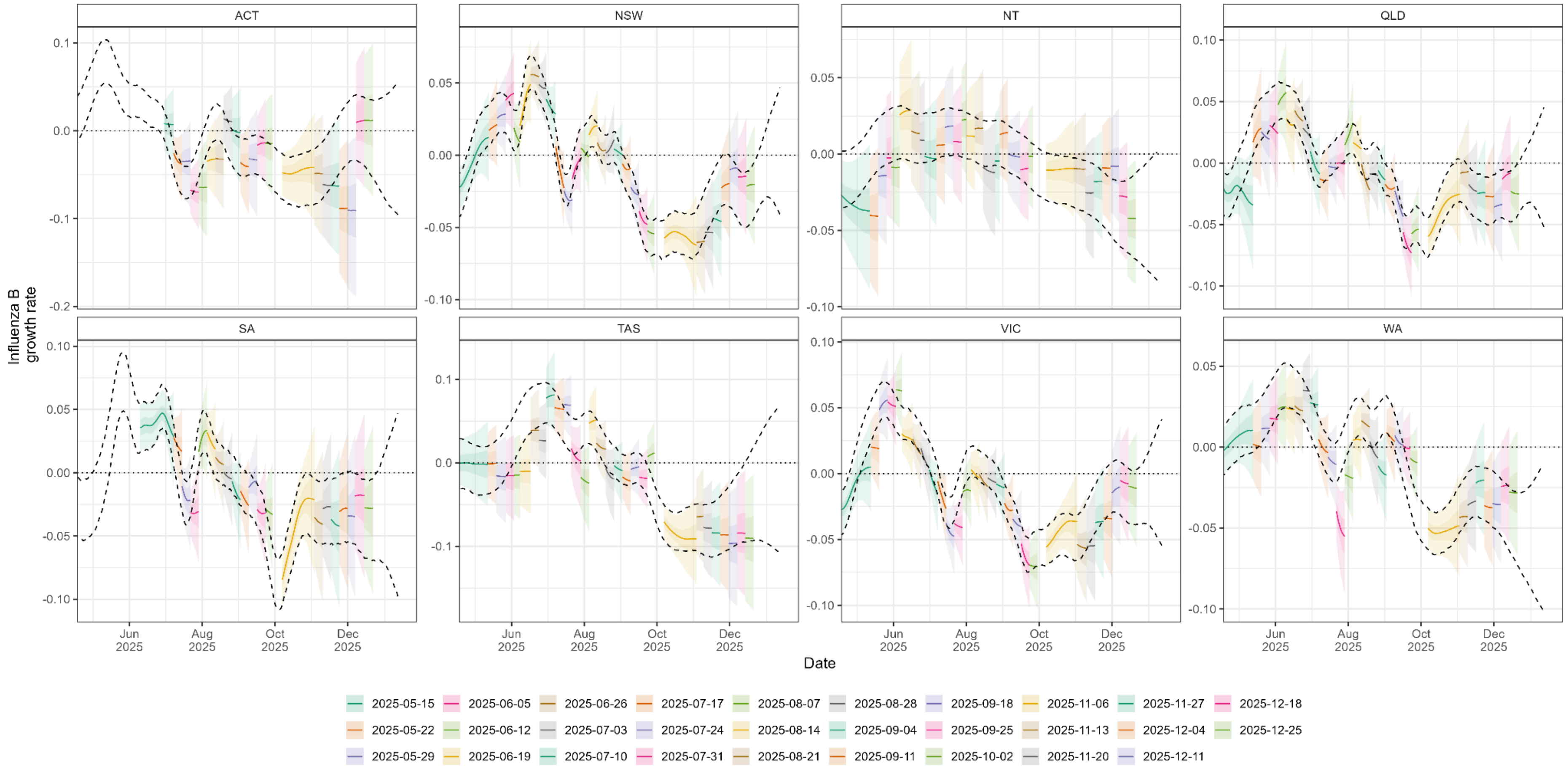


**Supplementary Figure 23: Real-time estimates of the growth rate of influenza B.** Growth rate (lines and shaded region) inferred from modelled trends in influenza B cases for all jurisdictions. Real-time estimates (coloured lines and shaded regions) are compared to 95% credible intervals of end-of-season estimates (black dashed lines). For all real-time estimates (coloured by round) we include the model posterior distribution's median (line) and 50% and 95% credible intervals (dark shaded and light shaded regions). We include the real-time model estimates up to the final day of data used when fitting the model (for periods greater than the previous round's model fit).

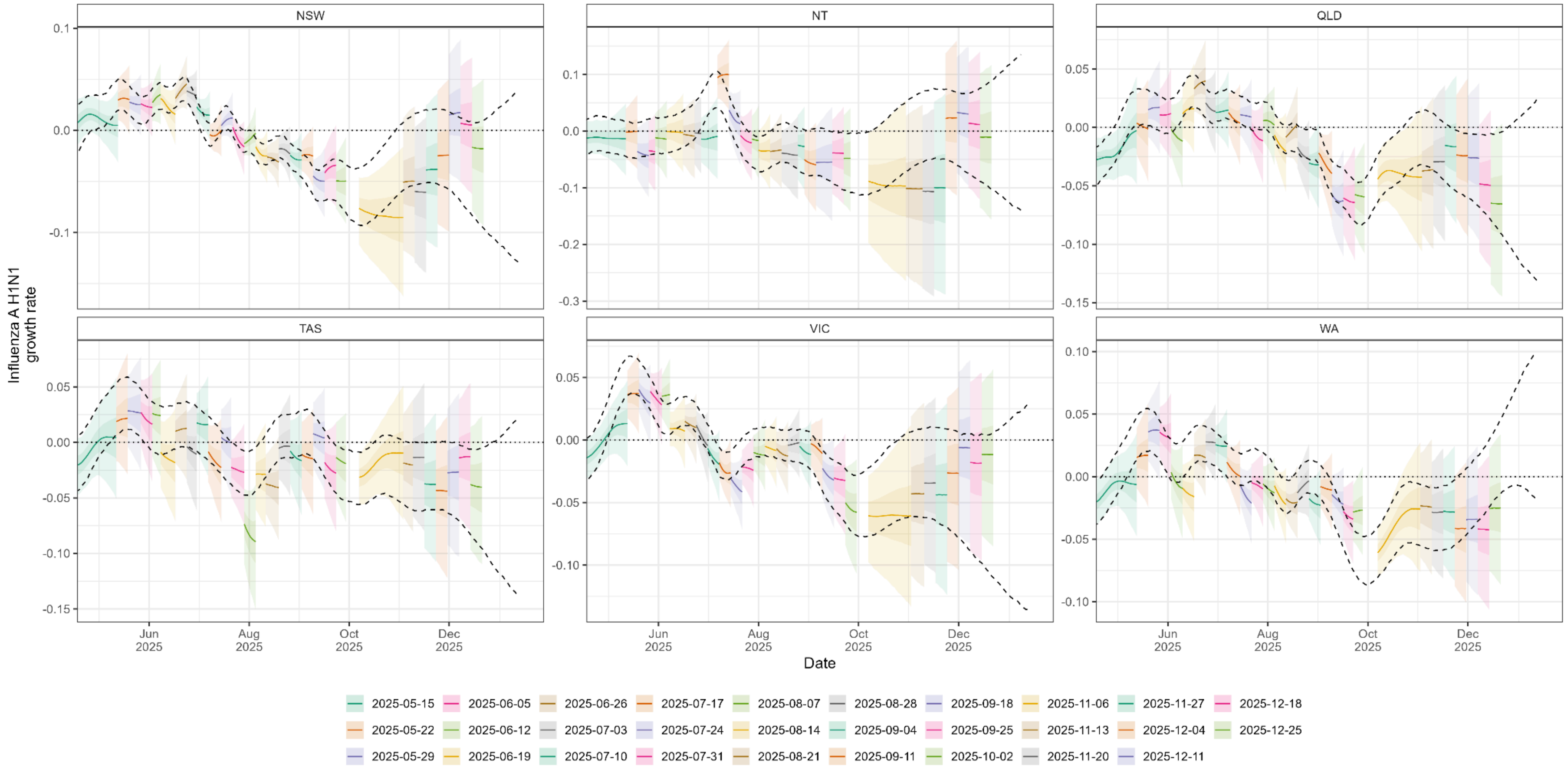


**Supplementary Figure 24: Real-time estimates of the growth rate of influenza A H1N1.** Growth rate (lines and shaded region) inferred from modelled trends in influenza A H1N1 cases for all jurisdictions where subtype data were available. Real-time estimates (coloured lines and shaded regions) are compared to 95% credible intervals of end-of-season estimates (black dashed lines). For all real-time estimates (coloured by round) we include the model posterior distribution's median (line) and 50% and 95% credible intervals (dark shaded and light shaded regions). We include the real-time model estimates up to the final day of data used when fitting the model (for periods greater than the previous round’s model fit).

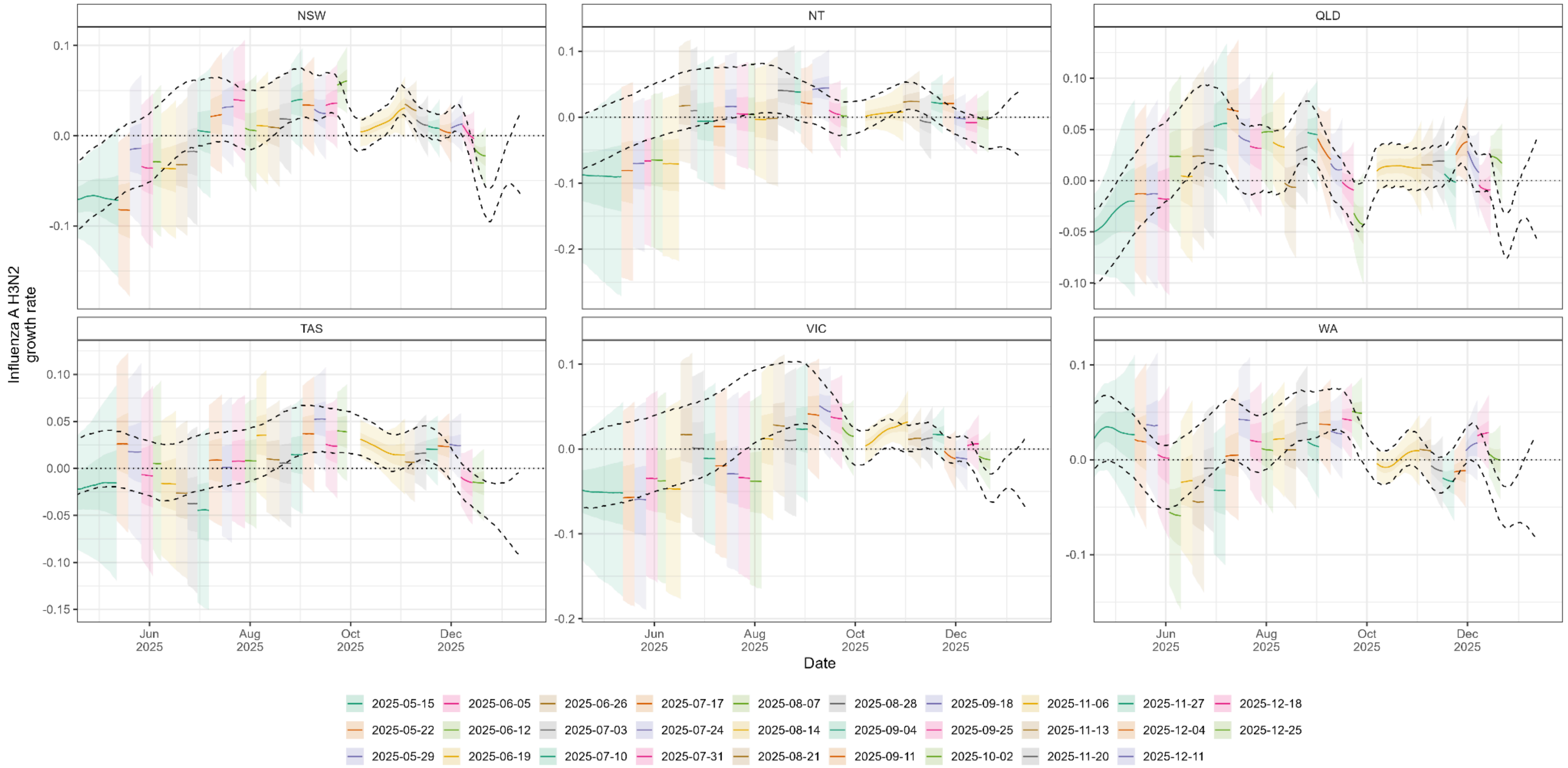


**Supplementary Figure 25: Real-time estimates of the growth rate of influenza A H3N2.** Growth rate (lines and shaded region) inferred from modelled trends in influenza A H3N2 cases for all jurisdictions where subtype data were available. Real-time estimates (coloured lines and shaded regions) are compared to 95% credible intervals of end-of-season estimates (black dashed lines). For all real-time estimates (coloured by round) we include the model posterior distribution's median (line) and 50% and 95% credible intervals (dark shaded and light shaded regions). We include the real-time model estimates up to the final day of data used when fitting the model (for periods greater than the previous round's model fit).

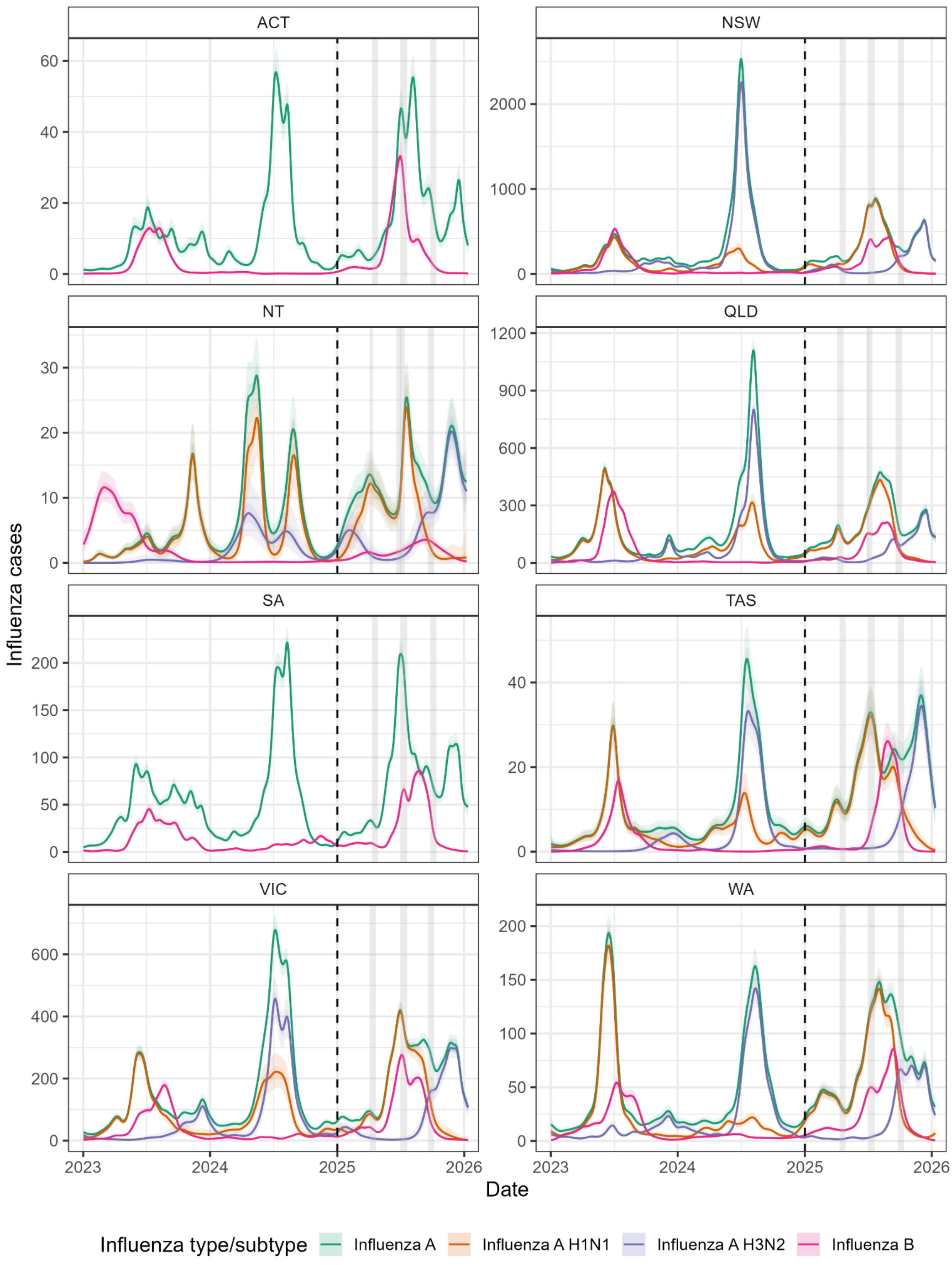


**Supplementary Figure 26: Modelled case dynamics of influenza subtypes over the previous three seasons with influenza A shown for all jurisdictions.** Modelled estimates of the expected number of cases over time for each influenza subtype. All estimates are shown over time with median (line), 50% credible intervals (dark shaded region) and 95% credible intervals (light shaded region). The vertical dashed line highlights the start of the 2025 season which is the focus of this paper. The grey shaded regions highlight the Autumn, Winter and Spring school holidays for each jurisdiction in 2025.
